# FGF21 reflects a responsive adipose tissue-liver axis in both cardiometabolic burden and following metabolic surgery

**DOI:** 10.1101/2024.05.31.24307065

**Authors:** Marie Patt, Isabel Karkossa, Laura Krieg, Lucas Massier, Kassem Makki, Shirin Tabei, Thomas Karlas, Arne Dietrich, Martin Gericke, Michael Stumvoll, Matthias Blüher, Martin von Bergen, Kristin Schubert, Peter Kovacs, Rima M. Chakaroun

## Abstract

**Objective:** This research aims to uncover the factors associated with circulating FGF21 levels in a cohort mimicking metabolic disease progression, examining its relationship with adipose tissue (AT) morphology and function. It also investigates FGF21 level changes post-metabolic surgery, predictive factors, and their links to metabolic adjustments.

**Design:** In this observational study, serum FGF21 was measured in 678 individuals cross-sectionally and longitudinally in 189 undergoing metabolic surgery. We explored links between FGF21, AT histology, cardiometabolic risk factors, weight loss, glucose metabolism changes using feature selection algorithms, univariate/multivariate models, and transcriptome/proteome network analyses in subcutaneous and visceral AT.

**Results:** FGF21 levels track closely with central adiposity, subclinical inflammation, insulin resistance, and cardiometabolic risk, with circulating leptin emerging as the top predictor. Visceral AT inflammation was associated with liver dysfunction and FGF21 elevation. Post-surgery, FGF21 peaked transitorily at 3 months and predicted fat mass loss at 12 months but not HOMA-IR improvements. Mediation analysis indicated an increased catabolic and AT-lipolytic state associated with higher liver enzyme and FGF21 levels (total effect 0.38, p<0.01; proportion mediation 32%, p<0.01). AT fibrosis was related to a blunted transitory FGF21 increase, and lower fat loss, and hence linked with a reduced surgical effect (FFA and visceral AT fibrosis: rho=-0.31, p=0.030; FFA and fat-mass loss: rho=0.17, p=0.020).

**Conclusion:** FGF21 reflects the liver’s metabolic response to AT characteristics in both central adiposity and after metabolic surgery, with its dynamics reflecting AT-liver crosstalk.

## Introduction

Metabolic surgery yields substantial, long-term weight loss and significantly improves remission rates of metabolic syndrome (MetS) and type 2 diabetes (T2D) [1]. Changes in cytokine and hormonal signatures both in obesity and following metabolic surgery highlight the need to investigate putative metabolic regulators.

Fibroblast growth factor 21 (FGF21) is one such regulator, predominantly expressed in the liver and to lesser extents in the brain and pancreas under basal conditions. It is mainly secreted by the liver under stress stimuli, like enhanced lipoprotein catabolism and lipolysis, in prolonged fasting and mitochondrial stress [2,3]. FGF21 was first identified in 2005 as an insulin sensitizer, promoting glucose uptake in adipocytes via GLUT-1 upregulation in cell culture. Subsequent research has clarified potential *in vivo* glucoregulatory actions stemming from the inhibition of hepatic glucose output, enhanced peripheral glucose disposal, and insulin sensitivity [4]. These metabolic effects seem to be, at least partly, mediated by the FGF receptor 1 (FGFR1) in white adipose tissue (WAT), governing whole-body energy homeostasis and suggesting relevant crosstalk between these major metabolic organs [5,6]. Clinically, FGF21 analogs reduced hepatic steatosis [7] and improved fasting glucose and whole-body insulin sensitivity in individuals with obesity and T2D [8], but their impact on glycemic control has been inconsistent across studies [9].

Counterintuitively, FGF21 levels are elevated in obesity, T2D, and MetS [10,11], and conflicting evidence suggests a decrease after weight loss, although this trend varies depending on the weight loss method and the timing of measurement. While a small study showed no significant FGF21 changes one-year post-surgery, Roux-en-Y gastric bypass (RYGB) reportedly increases FGF21 levels, in contrast to dieting, which tends to decrease them [3,12–14]. These observations contrast with reports of FGF21 elevations after several days of fasting [3]. Additionally, the observed positive correlation between changes in HOMA-IR and FGF21 levels following weight loss, albeit not always reproducible due to the limited sample sizes, has led researchers to propose a potential role for FGF21 in the resolution of T2D [12,13,15]. Importantly, it is unclear whether WAT characteristics are relevant to FGF21 levels both at under homeostatic metabolic conditions and following metabolic surgery.

Accordingly, we postulated that a) variations in circulating FGF21 levels link with the spectrum of metabolic disease severity and relate to AT-characteristics including histological features, transcriptomes and proteomes; b) post-metabolic surgery changes in FGF21 levels associate with alterations in metabolic health markers such as obesity status, T2D remission, and insulin resistance; c) FGF21 levels at baseline, linked AT-characteristics and FGF21-changes after surgery are predictive of the aforementioned metabolic outcomes.

## Methods

### Study participants

678 participants, including 101 without MetS and 18.5< BMI <25 kg/m^2^ (labeled nonMS) were recruited to the University Hospital of Leipzig, Germany (Table S1). The remaining participants had various degrees of metabolic impairment. Of these, 189 individuals in a longitudinal subgroup underwent metabolic surgery (RYGB, n=168; VSG, n=21) and were followed up at either 3- and/ or 12-months post-surgery (Table S2). Exclusion criteria were inflammatory disorders, chronic renal disease, coronary artery disease, and pregnancy/breastfeeding. Baseline data were collected two months before surgery. T2D resolution after 12 months was defined as HbA1c<5.7%, fasting glucose<5.6 mmol/l, and no antidiabetic medication.

Obesity was defined as a BMI≥30 kg/m^2^ (ICD-10-GM 2021 criteria). T2D was diagnosed using the American Diabetes Association (ADA) 2019 criteria [16] and assessed dichotomously, while other forms were excluded based on clinical presentation, c-peptide, and auto-antibodies detection. Nutritional intake was evaluated using Food Frequency Questionnaires (FFQs) based on the European Prospective Investigation of Cancer (EPIC)-Norfolk-FFQ [17].

Phenotyping encompassed clinical (physical exams, blood pressure, ECG), anthropometric (body weight, height, BMI, waist-to-hip ratio [WHR], body impedance analysis using BC-418 MA, Tanita, Japan), and biological/metabolic assessments. The study protocols were approved by the University of Leipzig’s ethics committee (applications 017-12-23012012 and 047-13-28012013 [18,19]), with all participants providing written informed consent. Insulin resistance was indirectly measured using HOMA-IR [20]. Percentage Total Weight Loss (%TWL) was calculated as ((initial weight - weight at 12 months) / initial weight) * 100. Weight loss response was categorized based on %TWL, with <20% indicating a poor response and ≥20% a good response [21].

### Biochemical analyses including FGF21 measurements

Blood samples, taken after overnight fasting, were centrifuged immediately and stored at - 80°C for subsequent analysis. Kidney and liver functions, along with blood metabolic markers, were evaluated using standard clinical laboratory methods. Levels of interleukin 6 (IL6) and FGF21 were measured using ELISA kits (Biovendor, Czech Republic), following the manufacturers’ instructions. Sample absorbance was read in duplicates at 450 nm and referenced at 630 nm using a Tecan sunrise instrument (TECAN GmbH, Germany). The intra-assay and inter-assay coefficient of variation (CV) were 2.7% and 14.1%, respectively.

### AT histology

During metabolic surgery, abdominal subcutaneous and visceral WAT (scAT and visAT) biopsies were collected from 59 individuals. Samples were either immediately frozen in liquid nitrogen and stored at −80°C or fixed in 4% paraformaldehyde at 4°C for 48 hours, then embedded in paraffin and sectioned into 6 µm slices. Adipocyte size and counts, macrophages, and crown-like structures were quantified as previously detailed [18]. ScAT fibrosis was assessed by staining for type I and III collagen using picrosirius-red, with the ratio of collagen to total tissue area indicating fibrosis levels [22]. Fibrosis was classified as perilobular (PLF) and pericellular (PCF), each scored from 0 (none or very limited) to 2 (severe). A combined semiquantitative fibrosis score (FAT-score) was assigned to each sample in a blinded fashion, ranging from 0 (no fibrosis) to 3 (severe PLF and PCF) [18].

### AT transcriptomics and proteomics

We examined transcriptomics and proteomics data [18] from scAT and visAT samples of the longitudinal subgroup (Table S3). The log2-transformed, normalized data from these tissues were screened for outliers as previously described [18]. Data free of outliers (excluding two scAT and visAT transcriptome samples and three scAT proteome samples) were used to assess Spearman’s rank correlations with baseline FGF21 levels, focusing on the top 25 positively and negatively significantly correlating transcripts or proteins (with ENSG IDs) (Tables S4-S7).

Significant protein-transcript overlaps were identified using Uniprot-to-ENSG ID mappings via DAVID[23]. Weighted Gene Correlation Network Analyses (WGCNAs) [18] were performed, with the following specific parameters (scAT transcriptome: maxBlockSize = 25000, soft-threshold = 7, module size = 500-5000; visAT transcriptome: soft-threshold = 5; scAT proteome: maxBlockSize = 5000, soft-threshold = 7, module size = 50-200). Signed networks yielded co-abundant protein or transcript modules. These modules were correlated with FGF21 levels, total fat mass loss, weight loss percentage, and T2D remission. Enrichment analyses identified affected biological processes using clusterProfiler and Gene Ontology Biological Processes (GO BPs) from MSigDB as previously described [18] (Tables S8-S11, 10.5281/zenodo.10402000). Significant terms (BH-adjusted p ≤0.0) were identified, and the top two enriched terms per module visualized.

### Statistics

Statistical analyses were conducted using R 3.6.1 and SPSS V.25. Mann-Whitney U test and the Wilcoxon signed-rank test for independent and paired group comparisons, respectively. Categorical data were analyzed using the Chi-square test. Non-parametric partial correlation was used to adjust correlations.

Feature selection was conducted using Boruta [24] to identify key variables influencing baseline FGF21 levels, which uses a random forest algorithm to establish the feature importance score by comparing its relevance to that of randomly generated features.

Stepwise linear regression with backward elimination identified independent predictors for post-surgery changes. The area under the curve (AUC) for T2D remission and weight loss response was calculated using pROC [25]. FGF21 change-score was calculated as the difference between 3-months and baseline-FGF21 levels and regression models as well as recursive feature elimination built with scaled-centered data after excluding variables with high variance inflation factor values. The same approach was used to calculate change scores of other variables. Samples with missing data were excluded from further analyses.

Causal mediation analysis was performed using the mediation package [26] with 500 nonparametric bootstrapping simulations. P values<0.05 were considered significant and where necessary corrected for multiple testing according to Benjamini Hochberg considering p_FDR<0.05 as significant.

## Results

### FGF21 is associated with cardiometabolic burden

We first aimed to probe the links between FGF21 levels and cardiometabolic disease. FGF21 emerged as a significant marker of cardiometabolic and cardiovascular risk, as evidenced by its correlation with the Framingham ASCVD 10-year risk score (rho = 0.39, p<0.001, Figure 1A). FGF21 levels also showed specific associations with individual cardiovascular risk factors and disease severity: Notably, elevated FGF21 levels reflected more closely increased central adiposity (WHR, visceral fat mass) rather than total fat mass, and increased insulin resistance, subclinical inflammation (leucocyte counts, high-sensitivity C-reactive protein [hsCRP]), liver enzymes, as well as blood pressure (Figure 1B). These correlations persisted after adjustments for T2D status, age, BMI, and estimated glomerular filtration ratio (eGFR) (Table S12). The parallel between FGF21 levels and the severity of metabolic disease severity was further supported by its association with microalbuminuria, as a marker of early metabolic kidney damage and the number of antidiabetic, antihypertensive, and lipid-lowering medications prescribed (Figure 1B, Table S13).

**Figure 1:**
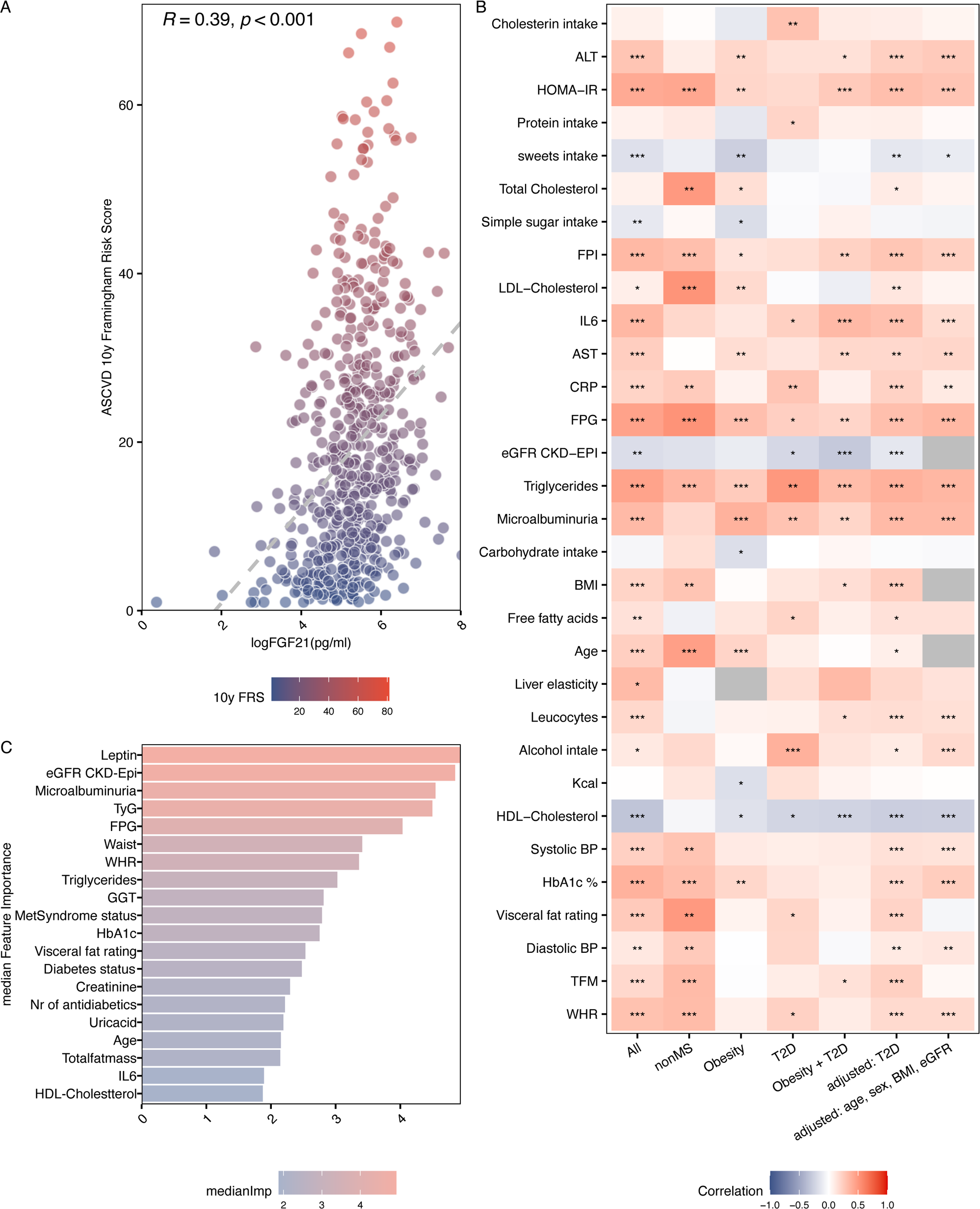
FGF21 along BMI and metabolic impairment spectrum. (**A**) Spearman’s ranked correlation between FGF21 serum levels and the Framingham ASCVD 10-year risk score (10y FRS). Point colors represent an escalating 10y FRS, transitioning from deep blue to a richer red. (**E**) Recursive feature elimination highlighting the main factors influencing FGF21 serum levels. Significance levels are denoted as: * for p≤0.05, ** for p≤0.01, and *** for p≤0.001. (**B**) Spearman’s ranked correlations illustrate the relationship between metabolic parameters and circulating FGF21. The direction of correlation is represented by color—positive (red) and negative (blue)—with the hue intensity denoting the magnitude of the correlation effect. (**C**) Feature importance in recursive feature elimination in the prediction circulating FGF21-levels. ***Abbreviations***: ALT (alanin aminotransferase), HOMA-IR (homeostatic model assessment of insulin resistance), FPI (fasting plasma insulin), IL6 (interleukin 6), AST (aspartate aminotransferase), CRP (c-reactive protein), FPG (fasting plasma glucose), eGFR (estimated glomerular filtration rate), BMI (body mass index), Kcal (estimated Kilocalories intake), HDL (high-density lipoprotein)., HbA1c (glycated hemoglobin), BP (blood pressure), TFM (total fat mass), TyG (triglyceride glucose index), WHR (waist-to-hip ratio), GGT (gamma-glutamyltransferase), MetSyndrome (metabolic syndrome).

To understand the relative importance of the many related features in predicting circulating FGF21, we performed recursive feature elimination on extensive clinical data and circulating proteins. This analysis highlighted kidney dysfunction, central adiposity, glucose control, and liver enzymes as key selected features. Notably, circulating leptin emerged as the top predictor, implying an association between FGF21 and AT, and highlighting its connection to metabolic and organ dysfunctions associated with central AT-distribution, including impairments in kidney and liver function (Figure 1C).

### Associations of circulating FGF21 with AT-traits reflecting lipid dysmetabolism and fibro-inflammatory pathways

As WAT and adipocytes are the main sources of leptin, the top predictor of FGF21 [27], we explored the connections between WAT traits and circulating FGF21 levels. Higher circulating FGF21 levels associated with increased adipocyte-size and reduced adipocyte-counts in both sc- and visAT. Elevated circulating FGF21 covaried with increased macrophage infiltration in visAT, which was uniquely correlated with increased liver GGT (Figure 2A).

**Figure 2:**
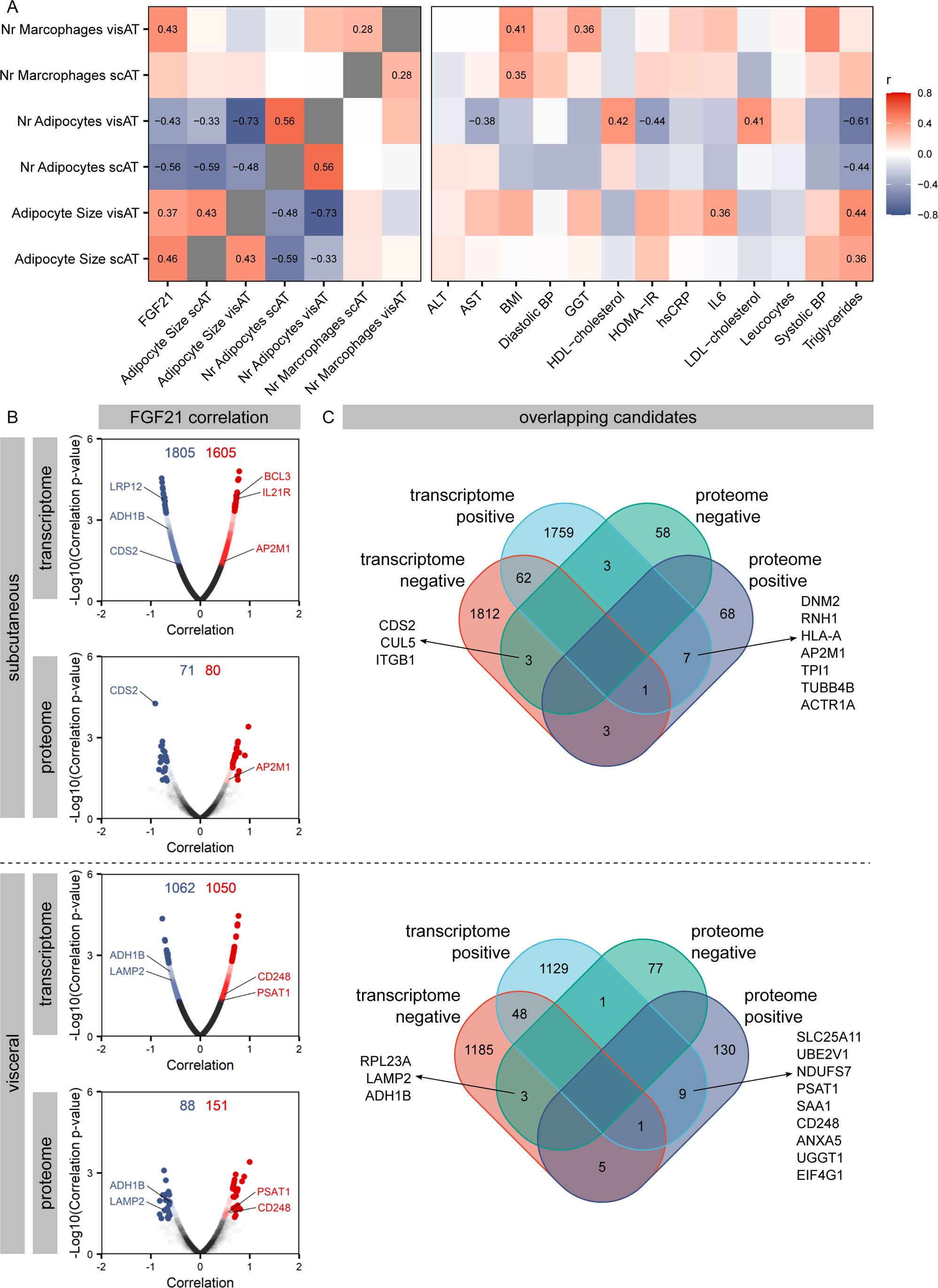
Correlation of FGF21 levels with AT characteristics. (**A**) Spearman correlations between circulating FGF21 levels and histological adipose tissue (AT) characteristics, alongside metabolic markers. Only tiles having significant correlations (p≤0.05) are labeled with their corresponding effect size. Correlation direction is denoted by color: positive correlations are red and negative ones are blue, with the depth of the hue reflecting the strength of the correlation. (**B**) Correlation assessments with circulating FGF21 were conducted for each transcript and protein derived from subcutaneous adipose tissue (scAT) or visceral adipose tissue (visAT). Candidates demonstrating significant positive correlations (p≤0.05) are presented in red, while those showing negative correlations are illustrated in blue. The cumulative count of these significant candidates is provided. Pertinent candidates are clearly marked. For clarity, the top 25 and bottom 25 candidates in terms of correlation strength are rendered opaque. (**C**) Venn diagram showcasing the overlap between transcripts and proteins that significantly correlate with circulating FGF21 either positively or negatively (termed transcriptome/proteome negative or positive). ***Abbreviations***: ALT (alanine aminotransferase), AST (aspartate aminotransferase), BMI (body mass index), BP (blood pressure), GGT (gamma glutamyltransferase), HOMA-IR (homeostatic model assessment of insulin resistance), CRP (c-reactive protein), IL6 (interleukin 6), LDL (low density lipoprotein), CDS2 (CDP-Diacylglycerol Synthase 2), CUL5 (Cullin-5), ITGB1 (Integrin Subunit Beta 1), DNM2 (Dynamin 2), RNH1 (Ribonuclease/angiogenin Inhibitor 1), AP2M1 (Adaptor Related Protein Complex 2 Subunit Mu 1), HLA-A (Major Histocompatibility Complex, Class I, A), TPI1 (Triosephosphate Isomerase 1), TUBB4B (Tubulin Beta 4B Class IVb), ACTR1A (ARP1 Actin-Related Protein 1 Homolog A, Centractin Alpha), RPL23A (Ribosomal Protein L23a), LAMP2 (Lysosomal Associated Membrane Protein 2), ADH1B (Alcohol Dehydrogenase 1B (Class I), Beta Polypeptide), SCL25A11 (Solute Carrier Family 25 Member 11), UBE2V1 (Ubiquitin Conjugating Enzyme E2 V1), NDUFS7 (NADH:Ubiquinone Oxidoreductase Core Subunit S7), PSAT1 (Phosphoserine Aminotransferase 1), SAA1 (Serum Amyloid A1), CD248 (Endosialin), ANXA5 (Annexin A5), UGGT1 (UDP-Glucose Glycoprotein Glucosyltransferase 1), EIF4G1 (Eukaryotic Translation Initiation Factor 4 Gamma 1)

Our analysis extended to specific transcripts and proteins within scAT and visAT, revealing a co-regulation with circulating FGF21 (Figure 2B, Figure S1). Key features consistently associated across both ATs included CDP-diacylglycerol synthase 2 (*CDS2*), BCL3 Transcription Coactivator (*BCL3)*[28], adaptor-related protein complex 2 subunit mu 1 (*AP2M1*)[29], and lysosomal associated membrane protein 2 (*LAMP2*) [30], implicated in lipid metabolism pathways (Figure 2C). Additional consistent features between transcriptome and proteome included alcohol dehydrogenase 1B (*ADH1B*), phosphoserine aminotransferase 1 (*PSAT1*), and *CD248* relating to fibro-inflammatory pathways.

Our analysis also revealed that in scAT, FGF21 associated with increased vesicle trafficking, intracellular transport, remodeling, reduced ubiquitin-proteasome pathway, and modified cell-cell interactions, as well as with lower *fibroblast growth factor receptor 1* (*FGFR1*) expression (rho=-0.44, p=0.04). In visAT, FGF21 correlated positively, with several transcripts and proteins relating to autophagy, phospholipid metabolism, and mitochondrial dysfunction, as well as with the marker of inflammation serum amyloid A1 (SAA1). However, no direct correlation was observed between FGF21 and AT-fibrosis, as indicated by the histological FAT-score in either AT (Table S14).

### FGF21 trajectory in metabolic surgery indicates liver catabolic response in the short term

Metabolic surgery is recognized for its impact on cardiometabolic improvement [3,9], despite existing debate concerning its impact on FGF21 [3]. We hence investigated FGF21 dynamics post-surgery. Overall, surgery led to substantial weight loss, improved insulin sensitivity, lipid profiles, and reduced inflammation, although with notable variability (Figure 3A, Table S2). Among 86 individuals with T2D at baseline, 57 achieved remission (Table S15).

**Figure 3:**
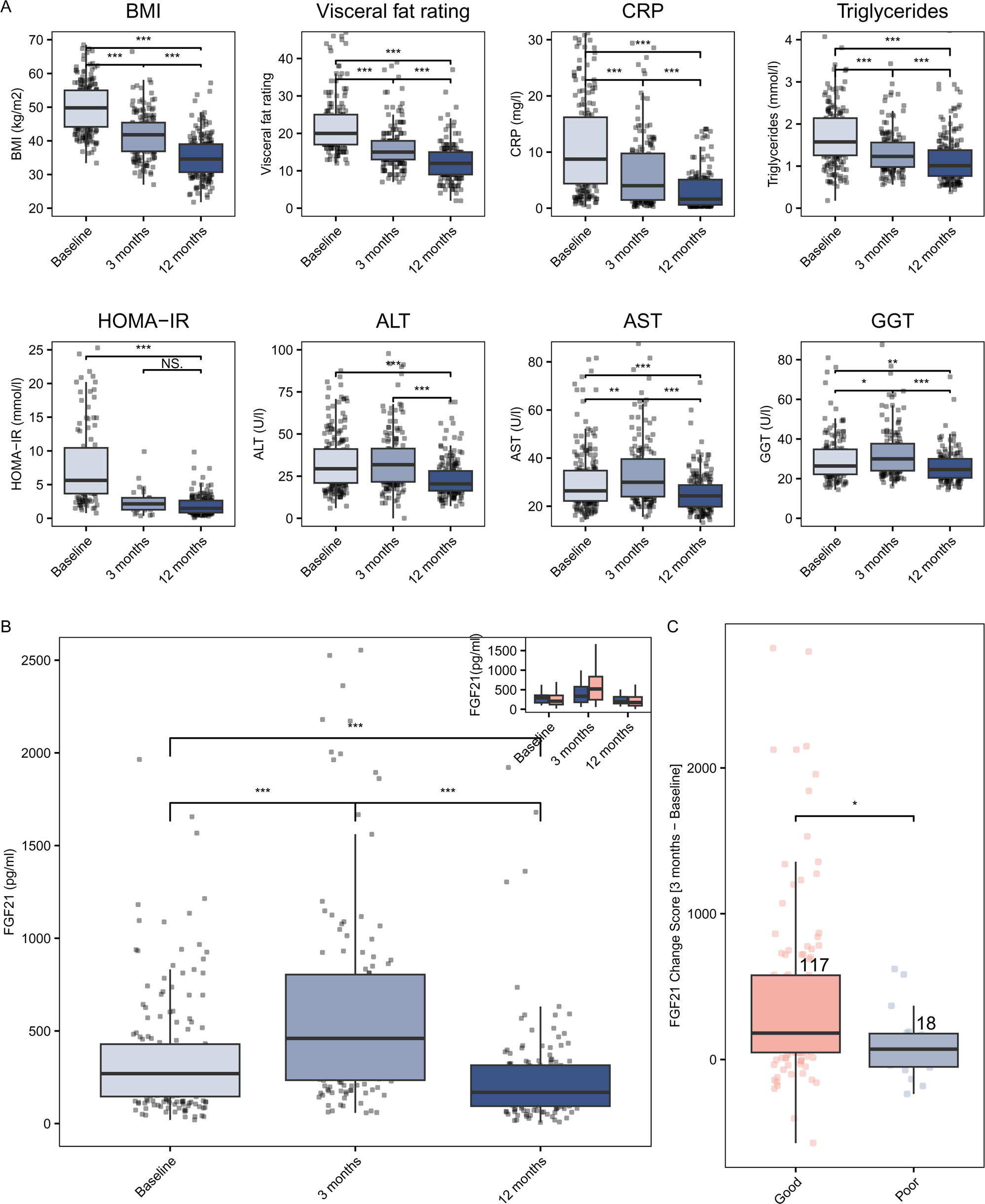
FGF21 dynamics following metabolic surgery. (A) Longitudinal changes following metabolic surgery in BMI (kg/m^2^), visceral fat rating, CRP (mg/l), triglycerides (mmol/l), HOMA-IR, ALT (U/l), AST (U/l), and GGT (U/l) at baseline, 3, and 12 months for up to 189 individuals. Statistical analyses: Mann-Whitney-U for overall differences; post-hoc Kruskal-Wallis test. For those with consistent data across timepoints (n=137), results were confirmed with Friedman’s ANOVA and Nemenyi post-hoc tests. (B) Longitudinal changes in FGF21 serum concentration (pg/ml) over the surgery using the same statistical tests as in (A). Inset boxplot differentiates results by response type: poor (blue, n=18) and good (light pink, n=122) responders. (**C**) Comparison of change score of FGF21 at 3 months between poor (n=18) and good responders (n=122) assessed using Kruskal-Wallis test. Significance is indicated by asterisks: *-p≤0.05, **-p≤0.01, ***-p≤0.001. ***Abbreviations***: BMI (body mass index), CRP (c-reactive protein), HOMA-IR (homeostatic model assessment of insulin resistance), ALT (alanine aminotransferase), AST (aspartate aminotransferase), GGT (gamma glutamyl transferase).

Circulating FGF21 levels initially increased at 3 months post-surgery, before declining below baseline levels at the 12-months mark (Figure 3B). The investigation of its short-term dynamics identified liver enzyme trajectories, and serum albumin as the only features significantly covarying with FGF21 change scores at 3-months post-surgery after correction for multiple testing. While FGF21 and liver enzymes changed in the same direction, higher increases of FGF21 at 3 months were correlated with lower serum albumin (Figures 3A, S4, Table S16). Importantly, sensitivity analyses reveal consistent results when the smaller VSG subgroup was excluded with a slightly reduced power for correlation analyses (Figure S5, Table S16).

### FGF21 dynamics predict weight loss but not improvement of insulin sensitivity at 12-months

Given the conflicting reports on the effects of metabolic surgery on FGF21 and its subsequent associations with metabolic improvements [3,15], along with the modest efficacy of FGF21 analogs in controlling glycemia [3], we explored the relationship between FGF21 and surgical outcomes: The reduction of FGF21 from baseline to 12 months paralleled improvements in metabolic parameters including glucose, lipid metabolism, and inflammation (Figure 3A, Table S13), with the 12-months CVD-risk inversely relating to the FGF21-decrease over 12-months (rho_framingham_= −0.190, p=0.021, Figure S3). However, post-surgery FGF21 levels maintained their positive association with CVD risk similar to baseline levels (rho_Framingham = 0.41, p < 0.001).

Accordingly, lower baseline FGF21 levels, as a reflection of overall better metabolic control, were linked to early insulin resistance improvements at 3 months, independent of weight loss (stepwise multivariate regression analysis, Table S17) with baseline FGF21 modestly predicting T2D remission at one year (AUC = 0.6, accuracy = 0.30).

However, the FGF21 change score at 3 or 12 months did not significantly predict HOMA-IR at 12 months in change score regression (with or without baseline correction) or recursive feature elimination (Table S18).

On the other hand, FGF21 dynamics significantly related to weight and fat loss outcomes: higher FGF21 change scores 3-months post-surgery were significantly higher in good responders (%TWL ≥20%) vs poor responders (%TWL <20%) (Figure 3C, S5 Table S19) and effectively differentiated between these two groups (AUC 0.73, accuracy of 0.66). Similarly, FGF21 change score at 3-months was a significant predictor for both weight and fat loss at one year as continuous variables (Tables S20, S21).

### ScAT architecture relates to FGF21 dynamics and T2D remission post-metabolic surgery

To decipher WAT processes linked to observed FGF21 changes post-metabolic surgery, we conducted WGCNAs on transcriptomes and proteomes from sc- and visATs, identifying modules of co-abundant transcripts and proteins relating to FGF21 levels post-surgery (Figure 4).

**Figure 4:**
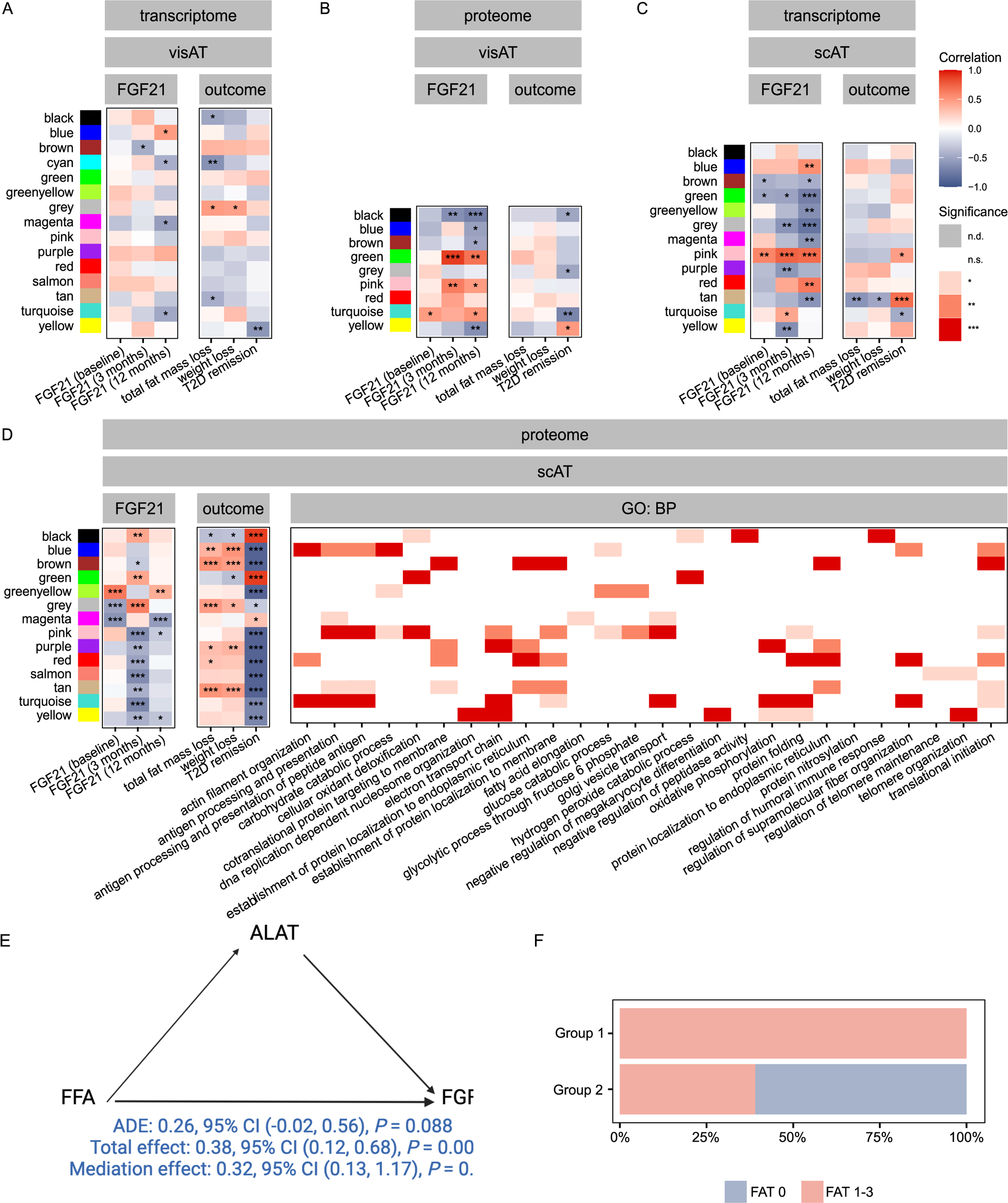
Correlation of clusters of transcripts and proteins from visAT and scAT with metabolic outcomes after metabolic surgery. WGCNA was applied to transcripts and proteins from baseline visAT (**A, B**) and scAT (**C, D**) The obtained modules of co-abundant transcripts and proteins were correlated with FGF21 levels and the selected metabolic outcomes. Correlation direction is denoted by color: positive correlations are red and negative ones are blue, with the depth of the hue reflecting the strength of the correlation. To get insights into the processes mirrored by the modules, enrichment analyses were conducted with all candidates assigned to one module. The top 2 enriched biological processes (BPs) from the Gene Ontology (GO) database are shown for the scAT proteome (**D, right panel**). The significance of enrichment is reflected by the intensity color. (**E**) Mediation analysis between FFA, ALAT as a marker of liver response and FGF21. The total effect of FFA on FGF21 (model including FFA and ALAT) was significant, with the average causal mediated effect (ACME) (the effect of the ALAT alone on FGF21) being significant, while the average direct effect and hence unmediated of FFA (ADE) was not significant. FFA influx hence only increases FGF21 when it elicits a response in the liver. Total effect = ADE + ACME. (**F**) Fraction of individuals with fibrosis (FAT score 1-3) and without fibrosis (FAT score 0) in scAT within individuals with <40% fat mass loss (group1, n=9) and individuals with ≥50% fat mass loss (group 2, n=25) (Chi-square analysis). Significance is indicated by asterisks: * - p≤0.05, ** - p≤0.01, *** - p≤0.001, NS. - not significant. *Abbreviations*: scAT (subcutaneous adipose tissue), visAT (visceral adipose tissue), FFA (free fatty acids), ALAT (Alanyl aminotransferase), ADE (average direct effect), ACME (average causal mediated effect), FAT score (fibrosis score of adipose tissue).

In the visAT transcriptome (Figure 4A), the yellow module showed a negative correlation with T2D remission, although no pathways were significantly enriched in this module. In contrast, three modules showed significant correlations with FGF21 levels and T2D remission in the visAT proteome (Figure 4B). Among these, the black module, which negatively correlated with FGF21 levels at both 3 and 12 months as well as with T2D remission, enriched for pathways involved in central carbon and fatty acid metabolism. Meanwhile, the turquoise module, alongside showing similar enrichments, also enriched for processes related to inflammatory responses and was related to higher FGF21 levels but reduced T2D-remission rates.

In the scAT transcriptome (Figure 4C), the tan module, related to growth and chemotactic processes (Table S8), positively associated with T2D remission, and negatively with 12-month FGF21 levels. Moreover, the scAT proteome (Figure 4D), showed most associations to FGF21 levels, including at 3-months, and T2D remission. In this analysis, modules related to immune responses and oxidant detoxification (green and black modules) showed a strong positive correlation with FGF21 levels at 3 months and T2D remission (rho=0.77, p=0.030), linking this desired outcome to the mentioned processes in scAT at baseline (Table S8).

### AT-liver axis mediates FGF21 levels in metabolic disease and surgery

We showed associations between FGF21 levels at baseline and 12 months mainly with visAT proteome and early post-surgery increase in FGF21 with scAT proteome. Also, the change in FGF21 short-term was linked mainly to increased markers of liver health and function, reduced albumin, and more importantly predicted fat mass loss. As a result, we wondered whether overlapping characteristics of insulin resistance and AT lipolysis were connected to FGF21 increases via the liver.

In this context, we explored whether free fatty acids (FFA), elevated due to AT insulin resistance and lipolysis [31] could serve as a putative link between visAT and hepatic production of FGF21. Causal mediation analysis revealed a significant effect of FFA on FGF21 levels (total effect 0.3, p=0.004). More importantly, the effect was dependent on liver response, as 32% of the total effect was accordingly mediated by the liver (p=0.004). More specifically, the mediator effect (ALAT) alone was significant (average causal mediated effect [ACME]: 0.12, p<2e-16), while the unmediated effect was statistically nonsignificant (average direct effect [ADE]: 0.12, p=0.088) (Figure 4E).

Since AT is the main source of FFA release, we explored whether FFA can be linked to AT-characteristics and therefore to metabolic outcomes. FFA levels were mainly positively associated with fat mass loss and negatively with visAT fibrosis scores (rho=0.17 and −0.31; p= 0.020 and 0.030, respectively). Accordingly, individuals with less than 40% fat mass loss had a more significant degree of fibrosis compared to those with fat mass loss of ≥50% (Figure 4F, Table S22). Importantly, we found a significant indirect effect of fat mass loss on FGF21 change scores at 3 months which was mediated by liver enzymes (ALAT, pACME = 0.028).

## Discussion

This study aimed to identify factors associated with serum FGF21 levels across a large cross-sectional cohort representing a range of metabolic disease severity. Additionally, it examined these levels in a longitudinal subset post-metabolic surgery, focusing on the relationship between FGF21-dynamics and metabolic improvements. We explored AT phenotypes, including histology, transcriptomics, and proteomics, to understand their potential interplay with FGF21 in surgical outcomes. Our research first confirms that elevated levels of FGF21 are linked with central adiposity, insulin resistance, systemic inflammation, and increased cardiometabolic risk [3,32]. Intriguingly, circulating leptin emerged as the top feature predicting circulating FGF21, suggesting a potential link between AT and FGF21 [27]. Indeed, we found a significant correlation between circulating FGF21 and adipocyte size in both sc- and visAT and accordingly with AT lipid storage (e.g., *CDS50* [33]). Importantly, we observed a unique association between circulating FGF21 with inflammatory SAA1 in visAT and with resident macrophages, which in turn, were linked to raised liver enzymes in our cohort. Accordingly, several features of visceral-adiposity-linked impairment such as the TyG-index as an indicator of insulin resistance reflecting glycemic control, along with circulating triglycerides [34], and elevated liver markers were among the top variables in the FGF21-prediction model.

Interestingly, despite the observed weight loss and metabolic improvements post-surgery, we noted an increase in FGF21 levels at the 3-month mark. This rise coincided with an increase in liver enzyme levels and a decrease in circulating albumin at the same time point. Additionally, liver enzymes were identified as highly important features in predicting FGF21 levels at baseline. Together, these results suggest that physiological changes linked to both insulin resistance in central adiposity, AT-lipolysis, and increased catabolism may disproportionally affect the liver as the primary link with FGF21.

VisAT fibrosis, on the other hand, shown to reduce lipolysis [35], was negatively linked to fat mass loss and FFA, suggesting that the increase in FGF21 at 3-months could be an indirect indicator of a responsive AT, with maintained capacity for lipolysis. Importantly, the link between FFA and FGF21 seemed to be entirely dependent on the liver’s response as indicated by mediation analysis. Similarly, the short-term increase in FGF21 also seems to link to immediate fat mass loss in a liver-dependent manner.

These results align with the understanding that FFAs, which increase due to the insufficient insulin-dependent inhibition of AT-lipolysis in states of insulin resistance, are strong determinants of FGF21 levels [3]. Furthermore, the acute rise in FFAs following metabolic surgery, leading to increased hepatic activation of PPAR-α [36], the key regulator of hepatic FGF21 [37], underscores the dynamic interplay between AT and liver response in the pathophysiology of obesity and weight loss.

The post-surgical increase in FGF21 could also relate to the central effects of food and protein restriction. This is partly supported by the negative correlation observed between caloric intake and FGF21 levels in our cohort, akin to FGF21 increases seen in extended fasting scenarios [3], although we lack post-surgery dietary intake data to fully support this hypothesis.

Considering recombinant FGF21’s effectiveness in reducing weight in rodent models of obesity [3,9,38], and mixed outcomes of FGF21 analogs on glycemic control [9,38], we investigated the relationship between FGF21 and metabolic as well as anthropometric outcomes following metabolic surgery. Our results indicate that the decrease in FGF21 levels is associated with an overall improvement in cardiometabolic risk. Additionally, the relationship between cardiometabolic burden and FGF21 levels at baseline and 12 months remains positive - higher cardiometabolic risk is correlated with elevated FGF21 levels both before and after metabolic surgery.

Interestingly, changes in the HOMA-IR score at one year were not directly related to adjustments in FGF21 levels at either 3 or 12 months. However, an initial lower FGF21 level predicted a short-term improvement in HOMA-IR, aligning with evidence that lower metabolic burden before surgery is associated with higher remission rates of T2D [39]. This suggests that improvements in HOMA-IR at 12 months may be largely secondary to weight loss.

Furthermore, changes in FGF21 levels at 3 months, were predictive of reductions in weight and fat mass at one year, in line with a recent adequately powered study [40]. Whether the initial increase in FGF21 contributes to fat mass loss and subsequent metabolic and inflammatory improvements at 12 months remains speculative. Supporting this speculation is the observation that increased PPAR-α activation enhances hepatic FA oxidation, contributes to fat depot reduction, and improves metabolic and inflammatory parameters and that FGF21 induces both lipolysis and AT-beigeing leading to increased energy expenditure [3,41]. Conversely, RYGB in FGF21-KO mice induced similar weight loss as in wild-type animals, suggesting FGF21 is not essential for weight loss after metabolic surgery [42].

### Limitations

Despite our unique integrative multi-omics approach providing numerous insightful observations of interest, our study is limited by the restricted sample size for AT proteomics and transcriptomics, and the lack of comprehensive diet data, which may add increased granularity for drawing conclusions on FGF21 dynamics post-surgery.

While we show a connection between AT and FGF21 via the liver, we do not know how much AT may contribute to circulating FGF21 after surgery. Prior to surgery, *FGF21*-expression in AT did not correlate with circulating FGF21 suggesting no direct contribution of AT to circulating FGF21 and in line with previous evidence of lack of very low FGF21-expression in scAT, visAT and epigastric AT [43]. Moreover, while we cannot exclude that FGF21 levels and their changes after surgery are due to FGF21 resistance in AT, *FGFR1*-expression correlated negatively with circulating FGF21 levels but only in scAT. As we do not have AT biopsies from 3- and 12-months, we are unable to investigate the effect of metabolic surgery on *FGFR1*-expression in adipose tissue and conclusively discuss the aspect of FGF21-resistance.

Importantly, our observations do not allow us to draw conclusions about causality or to pinpoint specific mechanisms.

Notwithstanding, our research uniquely integrates liver and adipose tissue data into a unified framework, revealing AT-liver crosstalk as a potential determinant of circulating FGF21 both in metabolic disease and after metabolic surgery. Additionally, we posit that visAT fibrosis can influence fatty acid availability and thus FGF21 secretion, adding a new dimension to our understanding of metabolic regulation, that merits further experimental investigation. Within this integrated framework, endogenous FGF21 reflects liver’s exposure to increased FFA: When visceral fat is increased, FFA are released through the portal system due to inadequate insulin-dependent inhibition of lipolysis leading to increased FGF21 in association with other markers of glucometabolic control. After surgery, healthier adipocytes, showcasing less fibrosis, are more lipolytic resulting in elevated levels of FFAs and FGF21, which are indicative of more favorable outcomes in weight loss, particularly when FGF21 concentrations are reduced owing to a decreased baseline rate of lipolysis after surgery. Within this specific context, short-term FGF21 change serves as a biomarker for healthier adipose tissue and a positive response to bariatric surgery, rather than acting as a primary effector of metabolic response as previously suggested by preclinical models, warranting further validation.

In summary, our study highlights FGF21’s maintained relationship with various cardiometabolic factors and its responsive adaptation to metabolic surgery. Crucially, our findings suggest that FGF21 levels may be influenced by hepatic FFA influx from AT with maintained lipolysis both in insulin resistance and following metabolic surgery, two sides of the “metabolic stress” coin.

## Data Availability

All data produced in the present study are available upon reasonable request to the authors

https://doi.org/10.5281/zenodo.10402000

## Acknowledgments

Microarray analysis was conducted at the Core Unit DNA Technology at the Faculty of Medicine of the University Leipzig, headed by PD Dr Knut Krohn, whom we thank. We thank Dr. Kassem Makki for critical discussions on the manuscript and Prof. Staffan Nilsson for statistical guidance. We thank Manuela Quandt, Beate Gutsmann and Steffi Ziesche for their excellent technical support and the study participants for their support. The graphical abstract was created using *Biorender.com*.

## Author Contributions

M.P., I.K., L.K., R.C. analysed data. M.P., I.K., R.C. wrote the manuscript. All authors reviewed and edited the manuscript.

## Guarantor’s statement

Dr. Rima Chakaroun is the guarantor of this work and, as such, had full access to all the data in the study and takes responsibility for the integrity of the data and the accuracy of the data analysis.

