## Supplementary material for "FGF21 reflects a responsive adipose tissue-liver axis in both cardiometabolic burden and following metabolic surgery": Table S1, Table S2, Table S3, Table S12, Table S13, Table S14, Table S15, Table S16, Table S17, Table S19, Table S22, Figure S1, Figure S2, Figure S3.

**Marie Patt^1^ *, Isabel Karkossa^2^ *, Ph.D., Laura Krieg^2^, Ph.D., Lucas Massier^1,3^, Ph.D., Kassem Makki^4^, Ph.D., Shirin Tabei^5,6^, M.Sc., Thomas Karlas^7^, Prof., Arne Dietrich^8^, M.D., Prof., Martin Gericke^9^, M.D., Prof. Michael Stumvoll^1,10^, M.D., Prof., Matthias Blüher^1,10^, M.D., Prof., Martin von Bergen^2,11,12^, Ph.D., Prof., Kristin Schubert^2^, Ph.D., Prof., Peter Kovacs^1,13^, Ph.D., Prof., & Rima M. Chakaroun^1,14^, M.D.**

1 University of Leipzig Medical Center, Medical Department III–Endocrinology, Nephrology, Rheumatology, Leipzig, Germany

2 Department of Molecular Systems Biology, Helmholtz-Centre for Environmental Research - UFZ, Leipzig, Germany

3 Department of Medicine (H7), Karolinska Institutet, Stockholm, Sweden

4 INSERM U1060, INRAE UMR1397, Université de Lyon

5 Institute of Endocrinology and Diabetes, University of Lübeck, Lübeck, Germany

6 Center of Brain, Behavior, and Metabolism (CBBM), University of Lübeck, Lübeck, Germany

7 University of Leipzig Medical Center, Medical Department II, Division of Gastroenterology, Leipzig, Germany

8 University of Leipzig Medical Center, Department of Visceral, Transplant, Thoracic and Vascular Surgery, Leipzig, Germany

9 Leipzig University, Institute of Anatomy, Leipzig, Germany

10 Helmholtz Institute for Metabolic Obesity and Vascular Research (HI-MAG), Helmholtz Zentrum München, University of Leipzig and University Hospital Leipzig, Leipzig, Germany

11 Institute of Biochemistry, Leipzig University, Leipzig, Germany

12 German Centre for Integrative Biodiversity Research (iDiv) Halle-Jena-Leipzig, Leipzig, Germany

13 Deutsches Zentrum für Diabetesforschung e.V., 85764 Neuherberg, Germany

14 Wallenberg Laboratory, Department of Molecular and Clinical Medicine and Sahlgrenska Center for Cardiovascular and Metabolic Research, University of Gothenburg, Gothenburg, Sweden

*** These authors contributed equally.**

**For Tables S18, 20, 21, Please Refer to: 10.5281/zenodo.10402000**

### **Table S1 Cross-sectional cohort characteristics grouped into nonMS, obesity, T2D, obesity + T2D**

|  | **Healthy** | | **T2D** | | **Obesity** | | **Obesity+T2D** | | p | n |
| --- | --- | --- | --- | --- | --- | --- | --- | --- | --- | --- |
| n | 101 | | 96 | | 247 | | 234 | |  | 678 |
| Sex (f/m) | 26/75 | bcd | 63/33 | acd | 99/148 | ab | 115/119 | ab | **6.5x10^-8^** | 678 |
| Age (years) | 51 [36; 64] | bcd | 67 [62; 71] | acd | 45 [34; 53] | abd | 60 [51; 65] | abc | **4.9x10^-47^** | 678 |
| FGF21 (pg/ml) | 87.9 [48.3; 178.1] | bcd | 222.5 [148.9; 421.0] | ac | 175.8 [101.7; 298.9] | abd | 289.3 [172.3; 488.2] | ac | **4.2x10^-25^** | 687 |
| BMI (kg/m^2^) | 22.6±2.0 | bcd | 27.8±1.6 | acd | 45.5±7.4 | abd | 41.5±9.5 | abc | **3.5x10^-96^** | 678 |
| WHR | 0.8±0.1 | bcd | 1.0±0.1 | acd | 0.9±0.1 | abd | 1.0±0.1 | abc | **6.2x10^-39^** | 630 |
| TFM (kg) | 16.3±5.8 | bcd | 24.9±5.7 | acd | 62.9±16.9 | abd | 52.8±21.2 | abc | **4.0x10^-92^** | 672 |
| Visceral fat rating | 5.5±3.1 | bcd | 13.0±2.9 | acd | 18.6±7.1 | ab | 18.8±7.0 | ab | **6.9x10^-64^** | 632 |
| Systolic BP (mmHg) | 118.7±14.6 | bcd | 136.9±13.5 | ac | 127.8±14.2 | abd | 132.7±15.1 | ac | **3.7x10^-18^** | 635 |
| Diastolic BP (mmHg) | 67.4±8.9 | bcd | 73.2±10.4 | a | 74.5±11.2 | a | 74.0±11.5 | a | **1.0x10^-7^** | 635 |
| FPG (mmol/l) | 4.9 [4.6; 5.3] | bd | 7.1 [6.1; 8.6] | ac | 5.3 [4.9; 5.7] | abd | 7.6 [6.2; 9.0] | ac | **1.3x10^-71^** | 674 |
| HbA1c (%) | 5.3 [5.2; 5.5] | bd | 6.5 [6.1; 7.1] | ac | 5.6 [5.4; 5.8] | abd | 6.7 [6.1; 7.3] | ac | **1.0x10^-85^** | 676 |
| FPI (mlU/l) | 4.8 [3.9; 6.5] | bcd | 10.4 [7.6; 16.3] | acd | 16.6 [11.3; 21.9] | ab | 18.6 [11.5; 32.7] | ab | **2.1x10^-50^** | 610 |
| HOMA-IR | 1.0 [0.8; 1.5] | bcd | 3.4 [2.2; 5.9] | ad | 3.9 [1.6; 5.4] | ad | 6.9 [3.5; 13.6] | abc | **4.9x10^-55^** | 610 |
| Total chol. (mmol/l) | 5.3±1.1 |  | 5.0±1.0 |  | 5.1±1.1 |  | 5.1±1.1 |  | 0.183 | 678 |
| HDL-chol.(mmol/l) | 1.9 [1.6; 2.2] | bcd | 1.3 [1.1; 1.6] | a | 1.3 [1.1; 1.5] | a | 1.2 [1.0; 1.4] | a | **5.5x10^-30^** | 678 |
| LDL-chol. (mmol/l) | 3.2±1.0 |  | 3.0±0.8 |  | 3.3±0.9 |  | 3.1±0.9 |  | 0.125 | 678 |
| Triglycerides (mmol/l) | 0.9 [0.6; 1.3] | bcd | 1.5 [1.2; 1.3] | a | 1.5 [1.1; 1.9] | ad | 1.7 [1.3; 2.3] | ac | **2.4x10^-27^** | 678 |
| eGFR CKD-EPI (ml/min per 1.73 m^2^) | 85.6 [77.6; 94.2] | c | 83.8 [74.6; 95.4] | c | 95.3 [82; 106] | abd | 86.3 [71.5; 100.5] | c | **1.9x10^-7^** | 677 |
| Microalbuminuria (mg/l) | 3.0 [3.0; 8.0] | bd | 8.5 [3.5; 24.0] | ac | 4.3 [3.0; 11.7] | bd | 9.5 [3.0; 30.5] | ac | **9.7x10^-9^** | 617 |
| AST (U/l) | 24.0 [20.4; 26.9] | bcd | 26.9 [22.8; 32.2] | ad | 25.7 [22.2; 32.3] | a | 30.5 [24.0; 38.6] | ab | **2.5x10^-9^** | 677 |
| ALT (U/l) | 17.4 [14.4; 22.2] | bcd | 26.9 [20.7; 35.2] | a | 26.9 [20.4; 38.9] | a | 31.2 [22.6; 45.5] | a | **2.3x10^-23^** | 677 |
| CRP (mg/l) | 0.7 [0.3; 1.5] | bcd | 2.2 [1.3; 4.6] | acd | 7.3 [3.5; 13.3] | ab | 5.4 [2.4; 11.9] | ab | **1.1x10^-44^** | 677 |
| IL6 (pg/ml) | 0.9 [0.6; 1.4] | bcd | 2.2 [1.5; 3.4] | acd | 2.9 [2.1; 4.4] | ab | 2.9 [2.1; 4.7] | ab | **2.0x10^-35^** | 529 |
| Leucocytes (x10^9^/l) | 5.2 [4.5; 6.4] | bcd | 6.4 [5.2; 7.6] | acd | 7.4 [6.3; 8.6] | ab | 7.3 [6.2; 8.9] | ab | **1.1x10^-23^** | 676 |
| Alcohol | 60.5 [17.5; 217.8] | cd | 47.1 [8.8; 126.1] | c | 9.45 [0; 50.4] | abd | 28 [0; 215] | ac | **2.4x10^-8^** | 482 |
| Caloric intake (kcal) | 1969.3 [1464; 2227.5] |  | 1795 [1388.9; 2248] |  | 1894.5 [1373; 2539.8] |  | 1891 [1418.8; 2186.7] |  | 0.665 | 482 |
| Carbohydrates | 233.8 [172.7; 337.3] |  | 206 [156.4; 257.3] |  | 210.9 [150.8; 324.5] |  | 207.2 [156.5; 253.1] |  | 0.196 | 482 |
| Cholesterol | 266.1 [172.7; 337.3] |  | 255.8 [183.3; 358.4] |  | 285.7 [201.5; 416.7] |  | 271 [202; 355.3] |  | 0.079 | 482 |
| Simple sugar | 104.5 [84.6; 133.7] | bd | 80.3 [55.6; 104.5] | ac | 91.8 [63.6; 129.6] | bd | 80.6 [58.5; 104.6] | ac | **4.9x10^-7^** | 482 |
| Sweets | 92.3 [58.6; 135] | bd | 72.3 [36.1; 103.4] | a | 71.3 [41.7; 118.2] |  | 62.4 [32.9; 104.8] | a | **0.001** | 482 |
| Protein | 73.1 [56; 90.9] |  | 77.7 [58.6; 97.1] |  | 82.2 [65; 101.7] |  | 77.5 [63.2; 97.2] |  | 0.084 | 482 |

All normally distributed values as mean ± standard deviation, others as median [25. percentile; 75. percentile]. Differences between groups were analyzed by Kruskal-Wallis ANOVA and between 2 individual groups with Mann-Whitney U test for independent groups. Differences in sex distribution was analyzed by x2 analysis. The Letter code indicates: (a) p < 0.05 vs healthy, (b) p < 0.05 vs T2D, (c) p < 0.05 vs obesity, (d) p < 0.05 vs obesity+T2D. ***Abbreviations***: BMI: body mass index, WHR: waist-to-hip ratio, TFM: total fat mass, BP: blood pressure, FPG: fasting plasma glucose, HbA1c: glycated hemoglobin, FPI: fasting plasma insulin, HOMA-IR: homeostatic model assessment of insulin resistance, HDL: high density lipoprotein, LDL: low density lipoprotein, eGFR CKD-EPI:  estimated glomerular filtration rate, AST: aspartate aminotransferase, ALT: alanine aminotransferase, CRP: c-reactive protein, IL6: interleukin 6

### **Table S2:** **Longitudinal cohort characteristics of individuals undergoing Roux-En-Y gastric bypass (RYGB) and vertical sleeve gastrectomy (VSG)**

|  | **RYGB** | | | | **VSG** | | | |
| --- | --- | --- | --- | --- | --- | --- | --- | --- |
|  | **Baseline** | **n** | **3 months** | **12 months** | **Baseline** | **n** | **3 months** | **12 months** |
| n | 168 | | | | 21 | | | |
| Sex (f/m) | 103/65 | | | | 09/12 | | | |
| Age (years) | 48.0±11.1 | | | | 44.9±10.8 | | | |
| FGF21 (pg/ml) | 270.3 [148.0; 440.4] | 168 | 465.5 [223.3; 812.0] | 175.1 [94.6; 312.8] | 271.1 [130.0; 364.1] | 21 | 461.2 [299.0; 742.7] | 164.4 [80.0; 323.0] |
| BMI (kg/m2) | 48.7±6.0 | 167 | 40.4±6.0 | 34.6±5.8 | 56.9±7.5 | 21 | 46.5±7.0 | 40.0±7.5 |
| TFM (kg) | 67.7 [57.2; 78.0] | 163 | 50.5 [39.7; 58.5] | 36.5 [26.5; 45.1] | 76.6 [63.1; 100.7] | 20 | 61.6 [51.4; 71.2] | 39.7 [29.7; 51.4] |
| Systolic BP (mmHg) | 131.8±15.1 | 131 | 119.0±14.1 | 119.2±15.1 | 130.2±12.0 | 20 | 114.2±16.5 | 117.2±10.1 |
| Diastolic BP (mmHg) | 76.1±12.6 | 131 | 68.3±10.9 | 69.1±10.9 | 68.7±10.4 | 20 | 62.6±8.7 | 65.7±8.3 |
| FPG (mmol/l) | 5.8 [5.2; 6.9] | 167 | 5.2 [4.8; 5.8] | 5.0 [4.6; 5.5] | 6.9 [5.5; 9.0] | 21 | 5.2 [5.0; 8.0] | 5.2 [4.9; 6.7] |
| HbA1c (%) | 5.8 [5.5; 6.5] | 166 | 5.3 [5.0; 5.7] | 5.2 [4.9; 5.5] | 6.2 [6.0; 7.3] | 21 | 5.8 [5.4; 6.8] | 5.5 [5.2; 6.2] |
| FPI (mlU/l) | 20.2 [12.9; 31.6] | 108 | 8.2 [6.6; 9.2] | 6.3 [3.9; 10.2] | 27.5 [22.9; 33.5] | 14 | 11.6 (mean) | 8.3 [4.6; 11.1] |
| HOMA-IR | 5.2 [3.3; 9.7] | 108 | 2.2 [1.3; 3.0] | 1.5 [0.8; 2.5] | 8.1 [6.5; 12.8] | 14 | 3.6 (mean) | 1.9 [1.0; 3.4] |
| Total cholesterol (mmol/l) | 5.0±1.1 | 168 | 4.1±0.9 | 4.2±0.7 | 4.6±1.0 | 21 | 4.5±1.0 | 4.7±1.0 |
| HDL-cholesterol (mmol/l) | 1.2±0.4 | 168 | 1.2±0.4 | 1.5±0.4 | 1.1±0.3 | 21 | 1.0±0.3 | 1.3±0.3 |
| LDL-cholesterol (mmol/l) | 3.1±1.0 | 168 | 2.4±0.7 | 2.4±0.6 | 2.9±0.9 | 21 | 2.9±0.9 | 2.9±0.9 |
| Triglycerides (mmol/l) | 1.5 [1.2; 2.0] | 168 | 1.2 [0.9; 1.5] | 1.0 [0.8; 1.3] | 1.6 [1.3; 2.6] | 21 | 1.5 [1.3; 1.9] | 1.1 [0.9; 1.5] |
| eGFR CKD-EPI (ml/min per 1.73 m2) | 91.3±23.5 | 167 | 94.8±19.1 | 95.0±18.8 | 99.8±20.9 | 21 | 98.1±22.5 | 99.3±23.7 |
| AST (U/l) | 27.0 [22.8; 34.8] | 167 | 30.0 [24.6; 39.3] | 25.2 [21.6; 30.6] | 22.8 [19.8; 31.2] | 21 | 25.8 [20.4; 34.8] | 19.8 [17.9; 25.0] |
| ALT (U/l) | 29.4 [21.3; 39.9] | 168 | 30.0 [21.6; 41.4] | 21.6 [17.4; 28.8] | 30.0 [22.8; 48.0] | 21 | 33.0 [17.4; 34.8] | 16.5 [13.4; 20.7] |
| CRP (mg/l) | 8.0 [4.0; 15.5] | 167 | 3.3 [1.3; 8.6] | 1.4 [0.6; 4.3] | 11.3 [6.8; 13.3] | 21 | 7.2 [4.8; 13.8] | 4.9 [1.6; 6.3] |
| Leucocytes (x109/l) | 8.1±2.2 | 168 | 7.2±1.9 | 6.7±1.7 | 8.8±2.7 | 21 | 8.3±2.6 | 7.1±2.0 |
| T2D status (yes/no) | 80/85 | 165 | 27/89 | 14/105 | 11/10 | 21 | 08/13 | 07/14 |
| TWL (%) |  |  | 17.5±4.2 | 29.5±7.8 |  |  | 18.2±4.4 | 29.8±10.1 |

All normally distributed variables as mean ± standard deviation, others as median [25. percentile; 75. percentile]. All individuals with FGF21 measurement at baseline were included. n: Number of individuals included. ***Abbreviations***: BMI: body mass index, WHR: waist-to-hip ratio, TFM: total fat mass, BP: blood pressure, FPG: fasting plasma glucose, HbA1c: glycated hemoglobin, FPI: fasting plasma insulin, HOMA-IR: homeostatic model assessment of insulin resistance, HDL: high-density lipoprotein, LDL: low-density lipoprotein, eGFR CKD-EPI:  estimated glomerular filtration rate, AST: aspartate aminotransferase, ALT: alanine aminotransferase, CRP: c-reactive protein, IL6: interleukin 6, TWL: total weight loss in %

### **Table S3: Subgroup characteristics with available transcriptomics and proteomics**

|  | **Mean/Median** | **n** |
| --- | --- | --- |
| Sex (f/m) | 29/30 | 59 |
| Age (years) | 48±11 | 59 |
| FGF21 baseline (pg/ml) | 387.4 [186.1; 709.0] | 58 |
| FGF21 after 3 months (pg/ml) | 385.6 [173.5; 765.1] | 24 |
| FGF21 after 12 months (pg/ml) | 190.1 [99.1; 311.0] | 24 |
| Weight baseline (kg) | 144.3±22.5 | 58 |
| Weight after 12 months (kg) | 99.0±16.3 | 53 |
| TFM baseline (kg) | 71.0±17.5 | 57 |
| TFM after 12 months (kg) | 35.7±12.4 | 50 |
| FPG baseline (mmol/l) | 6.0 [5.3; 7.1] | 59 |
| FPG after 12 months (mmol/l) | 5.0 [4.6; 5.7] | 53 |
| HDL-chol. baseline (mmol/l) | 1.1 [0.9; 1.4] | 59 |
| HDL-chol. after 12 months (mmol/l) | 1.5 [1.2; 1.7] | 53 |
| Triglycerides baseline (mmol/l) | 1.5 [1.2; 2.0] | 59 |
| Triglycerides after 12 months (mmol/l) | 1.0 [0.7; 1.6] | 53 |
| subcutaneous FAT-Score | 1 [0; 2] | 56 |
| visceral FAT-Score | 2 [1; 2] | 58 |
| T2D baseline (%) | 52.6 | 59 |
| T2D remission after 12 months (%) | 60.0 | 25 |

All normally distributed values as mean ± standard deviation, others as median [25. percentile; 75. percentile]. n: Number of individuals included in each analysis. ***Abbreviations***: TFM: Total Fat Mass, FPG: Fasting Plasma Glucose, FAT-Score: Fibrosis score of Adipose Tissue, T2D: Type 2 Diabetes

### **Table S12: FGF21 correlations with adjustment for T2D status, age, sex, BMI and eGFR**

|  | **T2D adjustment** | | | **Adjustment for age, sex, BMI, eGFR** | | |
| --- | --- | --- | --- | --- | --- | --- |
|  | **rho** | **p** | **Degrees of freedom** | **rho** | **p** | **Degrees of freedom** |
| BMI (kg/m^2^) | 0.24 | **4.1x10^-10^** | 675 |  |  |  |
| WHR | 0.18 | **3.0x10^-6^** | 627 | 0.18 | **7.0x10^-6^** | 623 |
| TFM (kg) | 0.23 | **1.7x10^-9^** | 669 | 0.04 | 0.335 | 665 |
| Visceral fat rating | 0.21 | **8.4x10^-8^** | 629 | -0.05 | 0.216 | 625 |
| Systolic BP (mmHg) | 0.14 | **2.8x10^-4^** | 632 | 0.13 | **8.3x10^-4^** | 628 |
| Diastolic BP (mmHg) | 0.09 | 0.018 | 632 | 0.11 | **0.005** | 628 |
| FPG (mmol/l) | 0.32 | **4.1x10^-17^** | 671 | 0.35 | **6.9x10^-20^** | 668 |
| HbA1c (%) | 0.19 | **7.7x10^-7^** | 673 | 0.24 | **1.7x10^-10^** | 670 |
| FPI (mlU/l) | 0.28 | **2.2x10^-12^** | 607 | 0.23 | **1.1x10^-8^** | 604 |
| HOMA-IR | 0.31 | **2.2x10^-15^** | 607 | 0.29 | **5.5x10^-13^** | 604 |
| Total cholesterol (mmol/l) | 1.00 | **0.011** | 675 | 0.05 | 0.185 | 671 |
| HDL-cholesterol (mmol/l) | -0.26 | **2.8x10^-12^** | 675 | -0.26 | **7.2x10^-12^** | 671 |
| LDL-cholesterol (mmol/l) | 0.12 | **0.002** | 675 | 0.05 | 0.176 | 671 |
| Triglycerides (mmol/l) | 0.37 | **6.8x10^-24^** | 675 | 0.345 | **3.1x10^-20^** | 671 |
| eGFR CKD-EPI (ml/min per 1.73 m^2^) | -0.14 | **3.9x10^-4^** | 674 |  |  |  |
| AST (U/l) | 0.18 | **2.0x10^-6^** | 674 | 0.19 | **6.8x10^-7^** | 671 |
| ALT (U/l) | 0.23 | **1.7x10^-9^** | 674 | 0.26 | **1.5x10^-11^** | 671 |
| CRP (mg/l) | 0.22 | **7.2x10^-9^** | 674 | 0.11 | **0.004** | 671 |
| IL6 (pg/ml) | 0.31 | **6.6x10-13** | 526 | 0.17 | **6.9x10^-5^** | 523 |
| Leucocytes (x10^9^/l) | 0.17 | **1.4x10^-5^** | 673 | 0.16 | **3.9x10^-5^** | 669 |
| Alcohol | 0.09 | 0.036 | 479 | 0.17 | **2.7x10^-4^** | 476 |
| Simple sugar | -0.05 | 0.246 | 479 | -0.07 | 0.128 | 476 |
| Sweets | -0.143 | **0.002** | 479 | -0.13 | **0.006** | 476 |

Non-parametric partial correlation was performed controlling 1. for T2D status and 2. for age, sex, BMI and eGFR. Only parameters with significant association resulting from spearman's rank correlation are listed. rho: correlation coefficients, p: associated P values. Bold values indicate statistically sig correlation at a value of p <0.05. n: Number of individuals included in each analysis. ***Abbreviations***: BMI: body mass index, WHR: waist-to-hip ratio, TFM: total fat mass, BP: blood pressure, FPG: fasting plasma glucose, HbA1c: glycated hemoglobin, FPI: fasting plasma insulin, HOMA-IR: homeostatic model assessment of insulin resistance, HDL: high density lipoprotein, LDL: low density lipoprotein, eGFR CKD-EPI:  estimated glomerular filtration rate, AST: aspartate aminotransferase, ALT: alanine aminotransferase, CRP: c-reactive protein, IL6: interleukin 6

### **Table S13: FGF21 and number of metabolic drugs at baseline and metabolic change after surgery**

| **Baseline** | **Serum FGF21 (pg/ml) baseline** | | |
| --- | --- | --- | --- |
|  | **rho** | **p** | **n** |
| # of antidiabtic drugs | 0.33 | **3.0x10-18** | 668 |
| # of antihypertensive drugs | 0.25 | **2.5x10-19** | 668 |
| # of lipid-reducing drugs | 0.22 | **1.2x10-8** | 662 |
| **Baseline to 12 months** | **Dynamic of serum FGF21** to 12months **(pg/ml)** | | |
|  | **rho** | **p** | **p adjusted** |
| FPG (mmol/l) | 0.340429 | **4.69E-05** | 7.02E-04 |
| HbA1c (%) | 0.174369 | **0.042327** | 0.153771 |
| HOMA-IR | 0.289216 | 0.260203 | 0.504235 |
| AST (U/l) | 0.321786 | **1.26E-04** | **0.001618** |
| GGT (U/l) | 0.319975 | **1.38E-04** | **0.001762** |
| ALT (U/l) | 0.383188 | **4.14E-06** | **7.58E-05** |
| Triglycerides (mmol/l) | 0.193441 | **0.02301** | 0.099443 |
| Systolic BP (mmHg) | 0.038204 | 0.671044 | 0.840334 |
| CRP (mg/l) | 0.021636 | 0.801847 | 0.911382 |
| Leukocytes (x10^9^/l) | -0.10335 | 0.227708 | 0.47033 |

Spearman’s rank correlation was performed. rho: correlation coefficients, p: associated P values. Bold values indicate statistically significant correlation at a value of p <0.05. n: Number of individuals included in each analysis. ***Abbreviations***: FPG: fasting plasma glucose, HbA1c: glycated hemoglobin, HOMA-IR: homeostatic model assessment of insulin resistance, AST: aspartate aminotransferase, ALT: alanine aminotransferase, GGT: gamma glutamyl transferase, BP: blood pressure, CRP: c-reactive protein

### **Table S14: Correlation of serum and WAT expressed FGF21 with WAT fibrosis**

| **visceral AT** | **Serum FGF21** | | | **FGF21 in visAT** | | |
| --- | --- | --- | --- | --- | --- | --- |
|  | **rho** | **p** | **n** | **rho** | **p** | **n** |
| PCF | 0.04 | 0.822 | 33 | 0.31 | 0.167 | 21 |
| PLF | 0.04 | 0.831 | 33 | 0.20 | 0.396 | 21 |
| FAT Score | 0.09 | 0.606 | 33 | 0.29 | 0.210 | 21 |
| **subcutaneous AT** | **Serum FGF21** | | | **FGF21 in scAT** | | |
|  | **rho** | **p** | **n** | **rho** | **p** | **n** |
| PCF | 0.11 | 0.556 | 32 | 0.30 | 0.184 | 21 |
| PLF | 0.05 | 0.792 | 32 | 0.18 | 0.443 | 21 |
| FAT Score | 0.11 | 0.557 | 32 | 0.22 | 0.345 | 21 |

Spearman’s rank correlation was performed. rho: correlation coefficients, p: associated P values. Bold values indicate statistically significant correlation at a value of p <0.05. n: Number of individuals included in each analysis. ***Abbreviations***: WAT: white adipose tissue, visAT: visceral adipose tissue, scAT: subcutaneous adipose tissue, PCF: pericellular fibrosis, PLF: perilobular fibrosis, FAT Score: fibrosis score of adipose tissue.

### **Table S15: Comparison of T2D remission and non-remission**

|  | **No T2D remission** | | | | | | **T2D remission** | | | | | | **Baseline** | | **3 months** | | **12 months** | |
| --- | --- | --- | --- | --- | --- | --- | --- | --- | --- | --- | --- | --- | --- | --- | --- | --- | --- | --- |
|  | **Baseline** | **n** | **3 months** | **n** | **12 months** | **n** | **Baseline** | **n** | **3 months** | **n** | **12 months** | **n** | **p** | **n** | **p** | **n** | **p** | **n** |
| n | 29 | | | | | | 57 | | | | | |  |  |  |  |  |  |
| Sex (f/m) | 10/19 | | | | | | 38/19 | | | | | | **0.004** | 86 |  |  |  |  |
| Age (years) | 50.0±12.2 | | | | | | 49.1±10.8 | | | | | | 0.602 | 86 |  |  |  |  |
| FGF21 (pg/ml) | 407.3 [208.9; 832.9] | 29 | 539.3 [243.3; 1099.7] | 20 | 257.8 [160.5; 441.9] | 21 | 314.8 [171.8; 451.9] | 57 | 435.4 [264.6; 927.5] | 49 | 219.5 [87.5; 363.2] | 49 | 0.109 | 86 | 0.968 | 69 | 0.135 | 70 |
| BMI (kg/m^2^) | 51.9±7.8 | 29 | 43.1±7.3 | 21 | 38.8±6.2 | 28 | 50.1±6.3 | 56 | 41.5±5.9 | 50 | 35.1±5.9 | 57 | 0.274 | 85 | 0.409 | 71 | **0.016** | 85 |
| TFM (in kg) | 73.9±20.9 | 27 | 54.0±20.8 | 21 | 42.6±15.8 | 26 | 70.8±17.8 | 56 | 52.0±15.8 | 50 | 37.6±15.6 | 57 | 0.502 | 83 | 0.743 | 71 | 0.190 | 83 |
| Visceral fat rating | 26.0 [20.0; 33.5] | 21 | 17.0 [15.5; 23.0] | 21 | 15.0 [13.0; 17.8] | 20 | 19.0 [17.0; 23.0] | 49 | 15.0 [13.0; 19.0] | 50 | 12.0 [8.0; 15.0] | 50 | **0.002** | 70 | 0.018 | 71 | **0.001** | 70 |
| FPG (mmol/l) | 8.9 [6.8; 12.0] | 28 | 7.6 [6.8; 9.4] | 21 | 7.1 [5.9; 8.9] | 29 | 6.9 [5.7; 8.1] | 57 | 5.6 [5.1; 6.5] | 50 | 5.1 [4.8; 5.7] | 57 | **0.006** | 85 | **2.5x10^-5^** | 71 | **2.0x10^-6^** | 86 |
| HbA1c (%) | 7.6 [7.1; 8.8] | 28 | 7.0 [6.6; 7.7] | 20 | 6.6 [5.5; 7.1] | 29 | 6.4 [5.8; 7.1] | 56 | 5.5 [5.3; 5.8] | 50 | 5.2 [5.0; 5.6] | 57 | **4.5x10^-5^** | 84 | **1.8x10^-7^** | 70 | **7.0x10^-7^** | 86 |
| FPI (mlU/l) | 28.6 [9.9; 38.5] | 18 | 12.8 [ ; ] | 2 | 9.3 [5.3; 13.2] | 25 | 28.0 [16.8; 54.1] | 31 | 8.6 [8.3; 15.4] | 8 | 9.2 [4.7; 10.6] | 46 | 0.507 | 49 | 1.0 | 10 | 0.357 | 71 |
| HOMA-IR | 11.4 [3.1; 17.2] | 18 | 4.5 [ ; ] | 2 | 3.1 [2.1; 4.3] | 25 | 10.2 [4.8; 18.7] | 31 | 2.8 [2.2; 4.3] | 8 | 2.1 [1.1; 2.7] | 46 | 0.724 | 49 | 0.400 | 10 | **0.005** | 71 |
| AST (U/l) | 30.9 [21.0; 36.0] | 28 | 28.2 [23.1; 36.9] | 21 | 21.6 [18.6; 27.6] | 29 | 31.8 [24.0; 42.9] | 57 | 31.2 [25.0; 42.4] | 50 | 24.6 [19.8; 32.4] | 57 | 0.308 | 85 | 0.249 | 71 | 0.120 | 86 |
| ALT (U/l) | 30.6±17.5 | 28 | 28.9±18.7 | 21 | 19.5±8.4 | 29 | 36.9±17.4 | 57 | 33.9±15.4 | 49 | 25.2±10.8 | 57 | 0.062 | 85 | 0.097 | 70 | **0.018** | 86 |
| CRP (mg/l) | 10.2 [5.6; 19.2] | 28 | 4.6 [2.5; 10.4] | 21 | 4.7 [1.1; 11.7] | 29 | 10.1 [5.0; 21.0] | 57 | 4.9 [1.7; 11.0] | 50 | 1.5 [0.7; 5.0] | 57 | 0.495 | 85 | 0.816 | 71 | **0.028** | 86 |
| Leucocytes (x10^9^/l) | 8.7±2.2 | 29 | 8.3±2.0 | 20 | 7.9±1.6 | 29 | 8.6±2.3 | 57 | 7.6±2.1 | 50 | 7.0±1.8 | 57 | 0.909 | 86 | 0.283 | 70 | **0.025** | 86 |
| Smoking (yes, %) | 27.8 | 29 | 10.0 | 21 | 22.2 | 29 | 27.8 | 57 | 14.3 | 50 | 21.9 | 57 | 1.0 | 54 | 0.731 | 38 | 0.977 | 50 |

All normally distributed variables as mean ± standard deviation, others as median [25. percentile; 75. percentile]. Only subjects with FGF21 measurement at baseline were included. Differences between no T2D-remission and T2D remission group for each time point were analyzed by Mann-Whitney-U test. Differences in fractions of sex, T2D and smoking status were compared by x2 analysis. p: associated P values. Bold values indicate statistically significant correlation at a value of p <0.05. n: Number of individuals included in each analysis. ***Abbreviations***: BMI: body mass index, TFM: total fat mass, FPG: fasting plasma glucose, HOMA-IR: homeostatic model assessment of insulin resistance, AST: aspartate aminotransferase, ALT: alanine aminotransferase, CRP: c-reactive protein.

### **Table S16: Associations of early post-surgery change scores with FGF21 change scores at 3 months**

| Chance scores of | **Spearman's Rho** | **nominal p-value** | **BH-adj p-value** | **Spearman's Rho RY**GB | **nominal p-value RYG**B | **BH-adj p-value RY**GB | **Status RY**GB |
| --- | --- | --- | --- | --- | --- | --- | --- |
| ALAT | 0.36 | 2×10^-05 | 0.001 | 0.38 | 3×10^-05 | 0.001 | cons. significant |
| ASAT | 0.32 | 0.00012 | 0.002 | 0.32 | 0.00042 | 0.008 | cons. significant |
| Serum albumin | -0.31 | 0.00027 | 0.003 | -0.31 | 0.00072 | 0.009 | cons. sig |
| GGT | 0.28 | 0.00106 | 0.01 | 0.29 | 0.00197 | 0.019 | cons. sig |
| microalbuminuria | 0.21 | 0.01708 | 0.1 | 0.24 | 0.01323 | 0.084 | cons. nonsig |
| TSH | 0.2 | 0.0185 | 0.1 | 0.23 | 0.01228 | 0.084 | cons. nonsig |
| HDL | -0.2 | 0.02135 | 0.101 | -0.18 | 0.0573 | 0.242 | cons. nonsig |
| TG | 0.18 | 0.03362 | 0.142 | 0.21 | 0.02228 | 0.121 | cons. nonsig |
| Total body water | 0.18 | 0.05482 | 0.208 | 0.17 | 0.08614 | 0.327 | cons. nonsig |
| HbA1c | 0.14 | 0.10536 | 0.364 | 0.12 | 0.21681 | 0.687 | cons. nonsig |
| eGFR | 0.12 | 0.17826 | 0.521 | 0.09 | 0.32663 | 0.875 | cons. nonsig |
| HSI | 0.1 | 0.25688 | 0.697 | 0.14 | 0.14915 | 0.515 | cons. nonsig |
| Uric acid | 0.09 | 0.3123 | 0.742 | 0.06 | 0.54753 | 0.91 | cons. nonsig |
| Muscle mass | 0.09 | 0.30153 | 0.742 | 0.08 | 0.41079 | 0.91 | cons. nonsig |
| HOMA-IR | -0.2 | 0.43356 | 0.845 | -0.08 | 0.77103 | 0.935 | cons. nonsig |
| insulin | -0.2 | 0.44495 | 0.845 | -0.04 | 0.87946 | 0.935 | cons. nonsig |
| weight | 0.07 | 0.39747 | 0.845 | 0.07 | 0.46211 | 0.91 | cons. nonsig |
| creatinine | -0.07 | 0.41856 | 0.845 | -0.03 | 0.7192 | 0.911 | cons. nonsig |
| Total fat free mass | 0.06 | 0.50525 | 0.914 | 0.04 | 0.66456 | 0.911 | cons. nonsig |
| Systolic blood pressure | 0.06 | 0.53966 | 0.926 | 0.08 | 0.39896 | 0.91 | cons. nonsig |
| Total cholesterol | 0.04 | 0.61257 | 0.926 | 0.04 | 0.64264 | 0.911 | cons. nonsig |
| Total fat mass | -0.05 | 0.56956 | 0.926 | -0.01 | 0.89531 | 0.935 | cons. nonsig |
| Total fat mass in % | 0.03 | 0.75575 | 0.926 | 0.05 | 0.59009 | 0.91 | cons. nonsig |
| Nr of antidiabetics | 0.04 | 0.67442 | 0.926 | 0.06 | 0.53388 | 0.91 | cons. nonsig |
| Nr of antihypertensives | -0.03 | 0.71392 | 0.926 | -0.04 | 0.71429 | 0.911 | cons. nonsig |
| Nr of antilipids | 0.03 | 0.70753 | 0.926 | 0.09 | 0.34559 | 0.875 | cons. nonsig |
| Hemoglobin | 0.04 | 0.66816 | 0.926 | 0.04 | 0.67179 | 0.911 | cons. nonsig |
| Visceral fat mass rating | -0.03 | 0.75475 | 0.926 | -0.01 | 0.92063 | 0.935 | cons. nonsig |
| Waist circumference | 0.05 | 0.59889 | 0.926 | 0.05 | 0.59839 | 0.91 | cons. nonsig |
| BMI | -0.02 | 0.78029 | 0.927 | -0.01 | 0.88952 | 0.935 | cons. nonsig |
| Diastolic blood pressure | 0.02 | 0.86635 | 0.968 | 0.06 | 0.55156 | 0.91 | cons. nonsig |
| CRP | 0.02 | 0.85552 | 0.968 | -0.06 | 0.55395 | 0.91 | cons. nonsig |
| LDL-cholesterol | 0 | 0.97463 | 0.975 | -0.02 | 0.79288 | 0.935 | cons. nonsig |
| sodium | 0.01 | 0.93984 | 0.975 | -0.01 | 0.93492 | 0.935 | cons. nonsig |
| leucocytes | 0 | 0.95955 | 0.975 | -0.02 | 0.81183 | 0.935 | cons. nonsig |
| platelets | -0.01 | 0.93909 | 0.975 | -0.07 | 0.47925 | 0.91 | cons. nonsig |

### **Table S17: Identification of predictors for insulin resistance (HOMA-IR) improvement 3 months after surgery**

| **Independent variables** | **Standardized β** | **SE** | **Sig.** | **R** | **Corrected r^2^** |
| --- | --- | --- | --- | --- | --- |
| Baseline FGF21 (pg/ml) | -0.666 | 0.003 | 0.011 | 0.734 | 0.441 |
| Baseline BMI (kg/m^2^) | -0.285 | 0.104 |  |  |  |
| %TWL | -0.140 | 0.114 |  |  |  |
| Baseline FGF21 (pg/ml) | -0.635 | 0.003 | 0.004 | 0.722 | 0.457 |
| Baseline BMI (kg/m^2^) | -0.272 | 0.102 |  |  |  |
| Baseline FGF21 (pg/ml) | -0.669 | 0.003 | 0.002 | 0.669 | 0.413 |

Linear regression analysis to assess predictors of HOMA-IR changes within 3 months after bariatric surgery. Independent variables evaluated: Fibroblastic growth factor 21 (FGF21), body mass index (BMI) at baseline and total weight loss in 12 months post-surgery (%TWL).

### **Table S19: Comparison of poor and good responders at baseline, 3 and 12 months post bariatric surgery**

|  | **Poor responder** | | | | | | **Good responder** | | | | | | **Baseline** | | **3 months** | | **12 months** | |
| --- | --- | --- | --- | --- | --- | --- | --- | --- | --- | --- | --- | --- | --- | --- | --- | --- | --- | --- |
|  | **Baseline** | **n** | **3 months** | **n** | **12 months** | **n** | **Baseline** | **n** | **3 months** | **n** | **12 months** | **n** | **p** | **n** | **p** | **n** | **p** | **n** |
| n | 19 | | | | | | 122 | | | | | |  |  |  |  |  |  |
| Sex (f/m) | 9/10 | | | | | | 89/33 | | | | | |  |  |  |  |  |  |
| Age (years) | 50.0±11.5 | | | | | | 47.2±10.9 | | | | | | 0.256 | 141 |  |  |  |  |
| FGF21 (pg/ml) | 279.2 [152.3; 359.7] | 18 | 299.0 [178.3; 578.3] | 19 | 179.9 [144.9; 323.0] | 18 | 209.5 [121.2; 357.5] | 117 | 520.5 [243.8; 841.4] | 117 | 171.4 [87.2; 318.4] | 117 | 0.334 | 141 | 0.060 | 136 | 0.422 | 139 |
| BMI (kg/m^2^) | 48.2±8.4 | 18 | 41.7±7.4 | 19 | 40.3±6.8 | 18 | 50.6±7.3 | 117 | 41.3±6.5 | 117 | 34.6±6.2 | 117 | 0.107 | 141 | 0.963 | 139 | **0.001** | 141 |
| Visceral fat rating | 23.0 [17.0; 34.0] | 18 | 17.0 [14.0; 27.0] | 19 | 15.0 [13.0; 21.0] | 18 | 20.0 [17.0; 25.0] | 117 | 15.0 [12.3; 18.0] | 117 | 10.0 [8.0; 14.0] | 117 | 0.282 | 140 | 0.057 | 139 | **4.5x10^-5^** | 139 |
| Systolic BP (mmHg) | 130.5±19.9 | 18 | 122.3±15.9 | 19 | 122.8±14.8 | 18 | 129.8±13.4 | 112 | 117.2±13.3 | 116 | 117.9±14.3 | 117 | 0.927 | 131 | 0.180 | 135 | 0.193 | 139 |
| Diastolic BP (mmHg) | 72.5±15.4 | 18 | 68.5±10.9 | 19 | 70.1±8.2 | 18 | 73.7±11.3 | 112 | 66.8±9.9 | 116 | 67.9±10.5 | 117 | 0.610 | 131 | 0.478 | 135 | 0.273 | 139 |
| FPG (mmol/l) | 7.3 [5.2; 9.2] | 18 | 5.4 [5.0; 7.6] | 19 | 6.1 [5.1; 7.2] | 18 | 5.8 [5.2; 6.9] | 117 | 5.2 [4.9; 6.0] | 117 | 5.0 [4.6; 5.3] | 117 | 0.070 | 140 | 0.188 | 139 | **3.6x10^-4^** | 141 |
| HbA1c (%) | 6.1 [5.7; 8.3] | 18 | 5.8 [5.5; 7.1] | 19 | 5.8 [5.1; 7.0] | 18 | 5.8 [5.5; 6.5] | 117 | 5.3 [5.0; 5.8] | 117 | 5.2 [4.9; 5.5] | 117 | 0.063 | 139 | **0.001** | 138 | **0.001** | 140 |
| FPI (mlU/l) | 25.2 [8.2; 35.3] | 8 | 12.8±7.1 | 2 | 9.9 [6.6; 12.4] | 15 | 21.4 [13.5; 32.4] | 72 | 8.2 [5.7; 9.2] | 22 | 5.7 [3.7; 10.6] | 95 | 0.898 | 80 | 0.406 | 24 | **0.015** | 110 |
| HOMA-IR | 7.7 [3.2; 12.2] | 8 | 4.5±2.1 | 2 | 2.8 [2.1; 3.3] | 15 | 5.9 [3.7; 13.1] | 72 | 2.0 [1.3; 2.6] | 22 | 1.5 [0.8; 2.5] | 95 | 0.688 | 80 | 0.087 | 24 | **0.001** | 110 |
| AST (U/l) | 23.4 [19.6; 35.1] | 18 | 25.2 [22.2; 30.6] | 19 | 20.4 [19.8; 25.2] | 18 | 26.7 [22.2; 34.9] | 122 | 30.6 [24.2; 39.0] | 120 | 24.6 [20.7; 30.6] | 121 | 0.228 | 140 | **0.024** | 139 | **0.030** | 140 |
| ALT (U/l) | 26.7 [14.8; 41.7] | 18 | 23.4 [15.5; 31.2] | 18 | 18.0 [15.0; 22.2] | 18 | 30.0 [20.8; 44.2] | 122 | 32.4 [21.6; 41.4] | 121 | 20.4 [16.8; 28.8] | 121 | 0.224 | 140 | **0.009** | 139 | 0.058 | 140 |
| CRP (mg/l) | 7.1 [4.6; 12.8] | 18 | 4.4 [2.2; 7.2] | 19 | 4.1 [0.9; 6.3] | 18 | 9.6 [4.4; 15.5] | 122 | 4.0 [1.4; 10.2] | 120 | 1.4 [0.6; 4.9] | 121 | 0.375 | 140 | 0.944 | 139 | 0.068 | 140 |
| Leucocytes (x10^9^/l) | 8.0±2.3 | 18 | 8.0±2.5 | 18 | 7.6±2.0 | 18 | 8.0±2.2 | 122 | 7.3±2.0 | 119 | 6.6±1.6 | 122 | 0.976 | 141 | 0.336 | 138 | 0.051 | 141 |
| Smoking (yes, %) | 30.0 | 10 | 9.1 | 11 | 9.1 | 11 | 14.9 | 122 | 9.7 | 122 | 11.4 | 122 | 0.235 | 77 | 0.717 | 73 | 0.819 | 81 |
| T2D status (yes, %) | 63.2 | 19 | 47.4 | 19 | 42.1 | 19 | 47.5 | 122 | 24.0 | 122 | 11.5 | 122 | 0.205 | 141 | **0.036** | 140 | **0.001** | 141 |
| T2D remission (yes, %) |  |  |  |  | 33.3 | 19 |  |  |  |  | 77.6 | 122 |  |  |  |  | **0.002** | 70 |

All normally distributed variables as mean ± standard deviation, others as median [25. percentile; 75. percentile]. Only subjects with FGF21 values at baseline, 3 and 12 months were included. Differences between poor and good responders for each time point were analyzed by Mann-Whitney-U test. Differences in fractions of sex, smoking status, T2D status and remission were compared by x2 analysis. p: associated P values. Bold values indicate statistically sig correlation at a value of p <0.05. n: Number of patients included in each analysis.

***Abbreviations***: BMI: body mass index, WHR: waist-to-hip ratio, TFM: total fat mass, BP: blood pressure, FPG: fasting plasma glucose, HbA1c: glycated hemoglobin, FPI: fasting plasma insulin, HOMA-IR: homeostatic model assessment of insulin resistance, AST: aspartate aminotransferase, ALT: alanin aminotransferase, CRP: c-reactive protein.

### **Table S22: Comparison of groups divided by fat mass loss**

|  | **Group 1 (<40% fat mass loss)** | | **Group 2 (≥50% fat mass loss)** | | **Baseline** | | **12 months** | |
| --- | --- | --- | --- | --- | --- | --- | --- | --- |
|  | **Baseline** | **12 months** | **Baseline** | **12 months** | **p** | **n** | **p** | **n** |
| n | 9 | | 25 | |  |  |  |  |
| Sex (f/m) | 4/5 | | 13/12 | | 0.697 | 34 |  |  |
| Age (years) | 56.1±7.5 | | 46.1±10.6 | | **0.014** | 34 |  |  |
| FGF21 (pg/ml) | 397.2 [102.4; 798.4] | 237.7 [88.8; 292.7] | 298.9 [162.7; 556.5] | 175.1 [97.2; 336.8] | 0.730 | 34 | 0.827 | 16 |
| BMI (kg/m^2^) | 50.8±7.0 | 38.5±4.2 | 48.1±7.1 | 31.8±3.8 | 0.188 | 34 | **0.001** | 33 |
| Visceral fat rating | 20.2±2.9 | 14.2 ±1.8 | 23.2±9.3 | 9.0±4.4 | 0.661 | 16 | **0.009** | 16 |
| FPG (mmol/l) | 6.5±1.2 | 7.8±5.5 | 6.7±2.3 | 5.1±0.8 | 0.514 | 34 | **0.011** | 34 |
| HbA1c (%) | 6.6±1.4 | 6.3±2.3 | 6.1±1.1 | 5.0±0.4 | 0.216 | 34 | **0.041** | 34 |
| HOMA-IR | 5.5±3.2 | 2.5±1.2 | 9.6±9.4 | 1.6±2.0 | 0.397 | 34 | **0.010** | 32 |
| AST (U/l) | 30.1±7.0 | 25.1±7.4 | 31.5±10.6 | 24.9±8.4 | 0.939 | 34 | 0.848 | 34 |
| ALT (U/l) | 28.1±11.8 | 20.3±7.5 | 35.0±13.1 | 24.2±11.1 | 0.163 | 34 | 0.397 | 34 |
| CRP (mg/l) | 17.8±12.2 | 9.0±11.0 | 12.2±12.7 | 2.7±3.8 | 0.163 | 34 | **0.024** | 34 |
| Leucocytes (x10^9^/l) | 8.2±1.1 | 7.5±1.2 | 8.2±2.2 | 6.6±1.6 | 0.730 | 34 | 0.151 | 34 |
| T2D status (yes, %) | 77.8 | 33.3 | 48.0 | 13.6 | 0.123 | 34 | 0.208 | 31 |
| T2D remission (yes, %) |  | 57.1 |  | 66.7 |  |  | 0.696 | 16 |

All variables as mean ± standard deviation. Except FGF21 as median [25. percentile; 75. percentile] due to high variability in circulating FGF21 levels. Differences between group 1 and group 2 for each time point were analyzed by Mann-Whitney-U test. Differences in fractions of sex, T2D status and remission of T2D were compared by x2 analysis. p: associated P values. Bold values indicate statistically sig correlation at a value of p <0.05. n: Number of individuals included in each analysis. ***Abbreviations***: BMI: body mass index, FPG: fasting plasma glucose, HbA1c: glycated hemoglobin, HOMA-IR: homeostatic model assessment of insulin resistance, AST: aspartate aminotransferase, ALT: alanine aminotransferase, CRP: c-reactive protein, T2D: Type 2 diabetes.

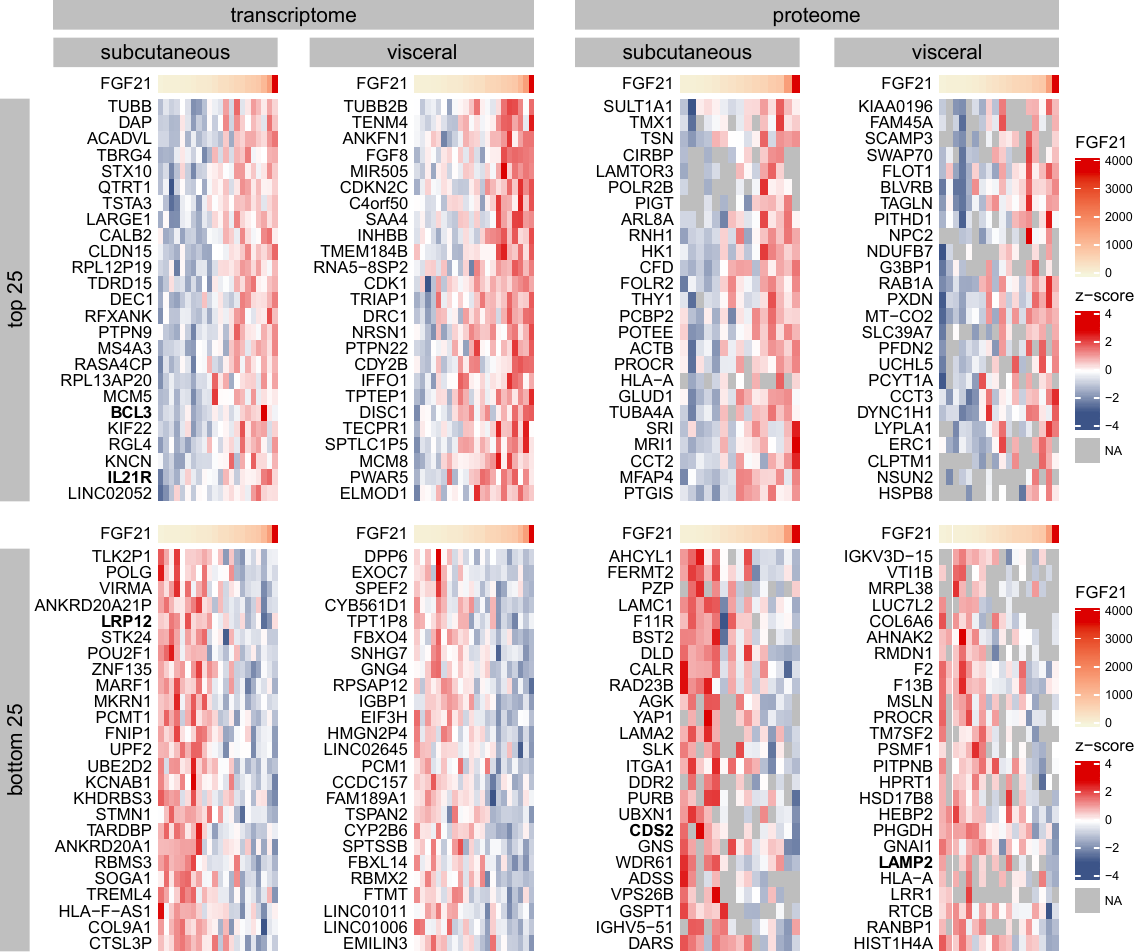

### **Figure S1: Spearman correlations between transcripts and proteins with baseline FGF21 levels.**

Shown are z-scored intensity data of the top and bottom 25 transcripts and proteins correlating sigly with baseline FGF21 levels (provided in pg/ml, Spearman correlation) in scAT and visAT. Relevant candidates are bold. Correlation direction is denoted by color: positive correlations are red and negative ones are blue, with the depth of the hue reflecting the strength of the correlation.

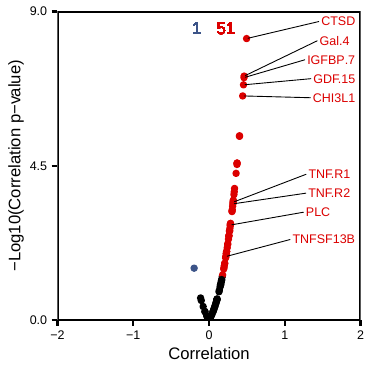

**Figure S2: Spearman correlations between FGF21 levels and circulating proteins at baseline.** Effect sizes are plotted against the log of the nominal p-value. Correlation direction is denoted by color: positive correlations are red and negative ones are blue. The number of sig positive and negative correlations left after correction for multiple testing are shown at the top (n=51 nad 1 respectively). Relevant candidates discussed in the text are labelled. ***Abbreviations***: CTSD (Cathepsin D), GAL4 (Galectin 4), IGFBP7 (Insulin-like Growth Factor-Binding Protein 7), GDF15 (Growth Differentiation Factor 15), CHI3L1 (Chitinase-3-Like Protein 1), TNFR1,2 (Tumor Necrosis Factor Receptor 1,2), PLC (Phospholipase C), TNFSF13B (Tumor Necrosis Factor Superfamily Member 13B)

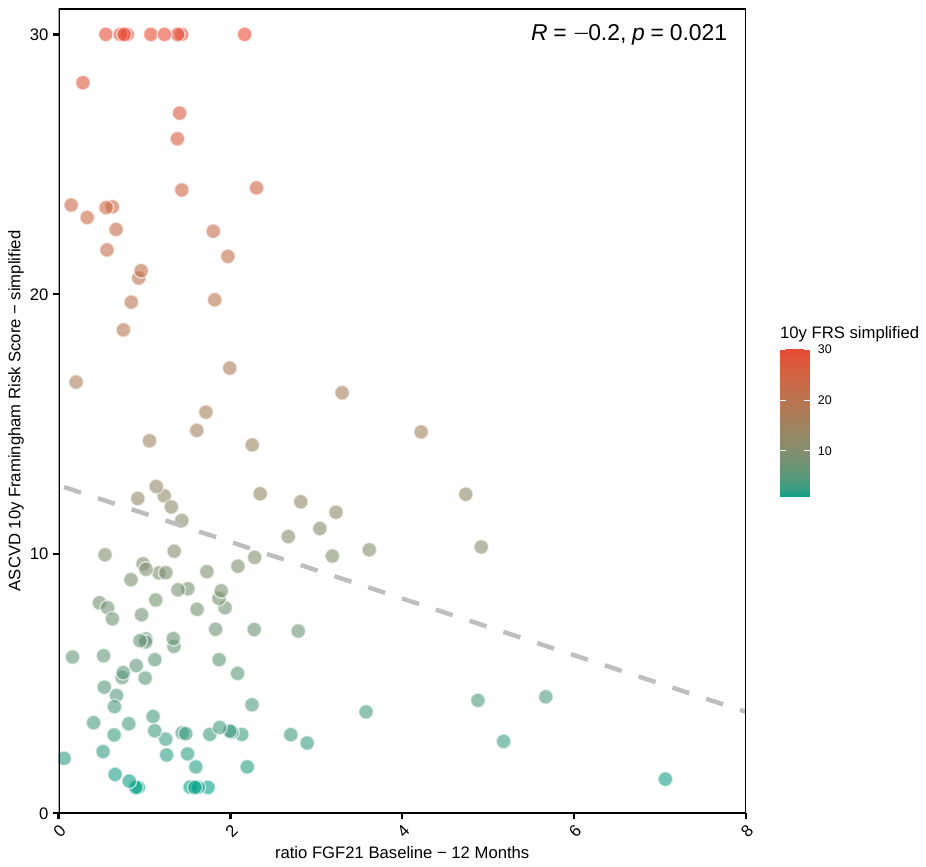

**Figure S3: Spearman’s correlation between ASCVD 10y Framingham Risk Score and the Ratio between FGF21 at baseline to FGF21 at 12 months.** The colour intensity from green to light red denotes an increased Risk Score.

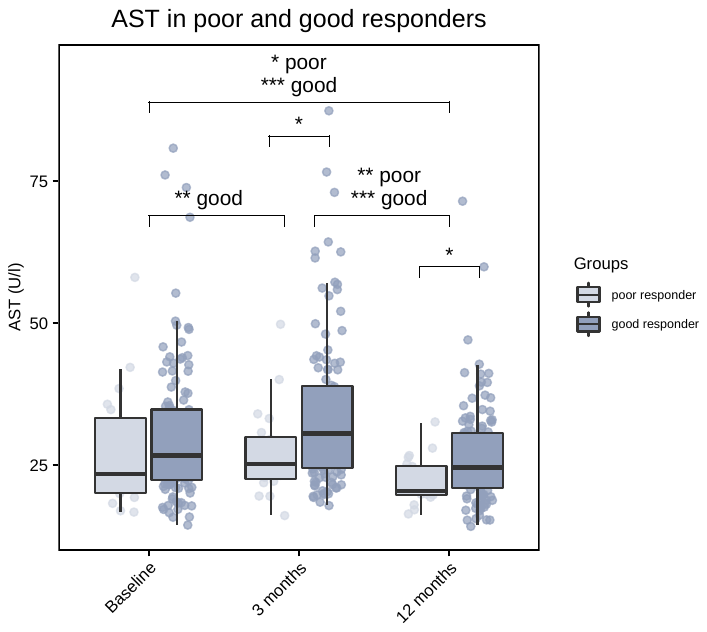

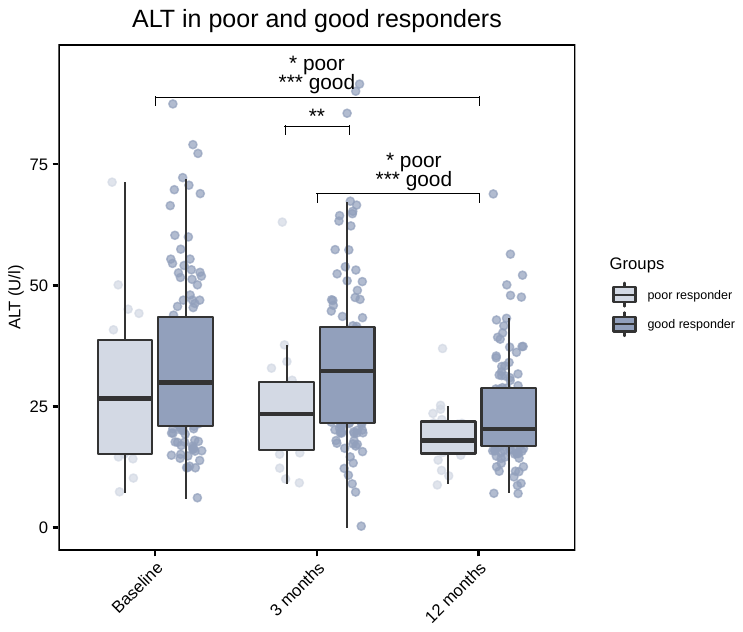

### **Figure S4: Change of AST and ALT serum levels in poor and good responders after surgery**

Dynamic of AST serum concentration (U/l) at baseline, 3 and 12 months after metabolic surgery in poor (n=18) and good responders (n=121) (Friedman’s ANOVA and post-hoc pairwise Nemenyi test). Poor responders are shown in light blue and good responders in dark blue. Significance is indicated by asterisks: * - p-value ≤0.05, ** - p-value ≤0.01, *** - p-value ≤0.001.

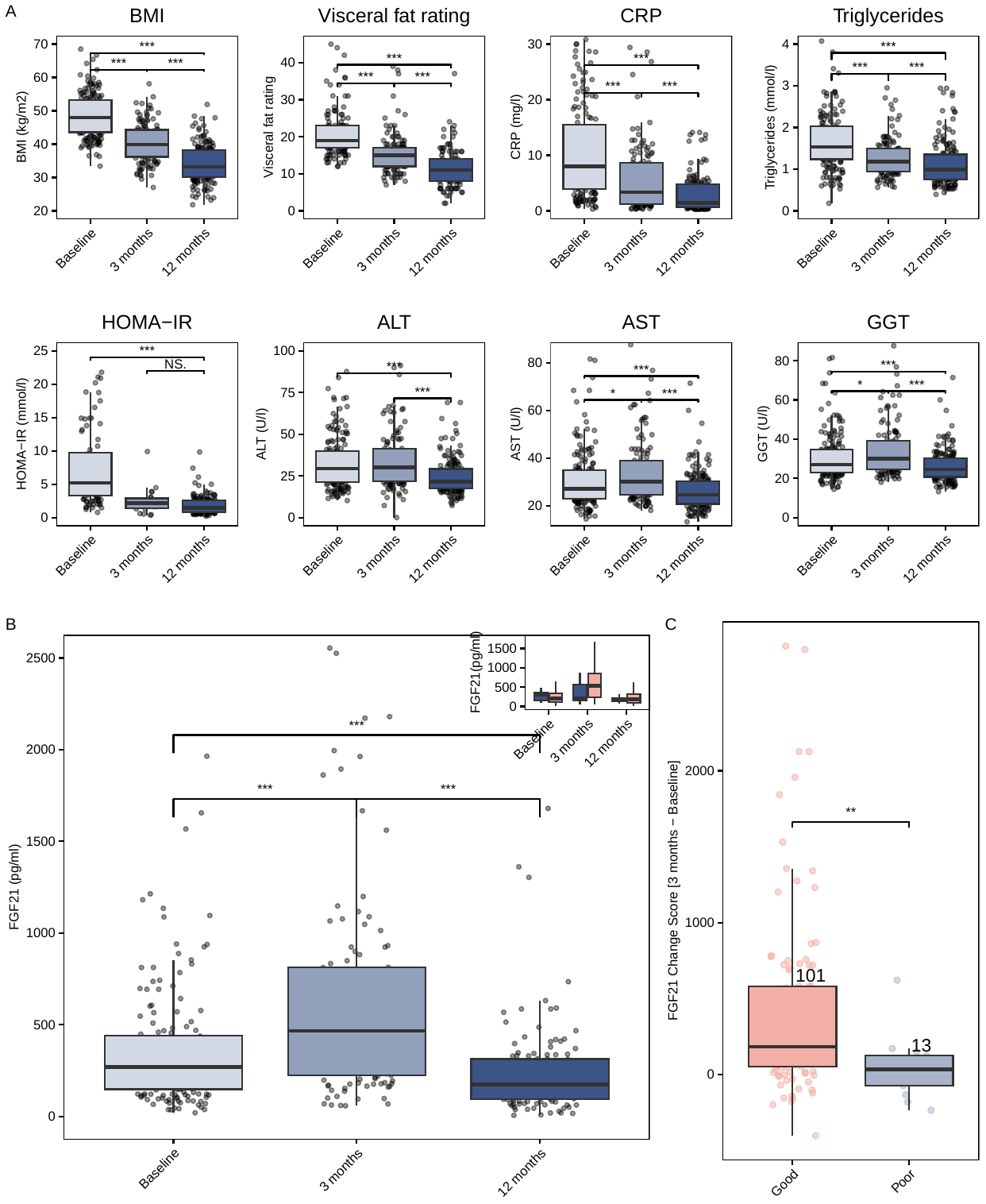

**Figure S5**: **FGF21** **dynamics following RYGB** (**A**) Longitudinal changes following metabolic surgery in BMI (kg/m^2^), visceral fat rating, CRP (mg/l), triglycerides (mmol/l), HOMA-IR, ALT (U/l), AST (U/l), and GGT (U/l) at baseline, 3, and 12 months restricted to the RYGB-subgroup.(**B**) Longitudinal changes in FGF21 serum concentration (pg/ml). Inset boxplot differentiates results by response type: poor (blue, n=13) and good (light pink, n=101) responders. (**C**) Comparison of FGF21 change score to 3 months between poor (n=13) and good responders (n=101) assessed using Kruskal-Wallis test. Significance is indicated by asterisks: *-p≤0.05, **-p≤0.01, ***-p≤0.001. ***Abbreviations***: BMI (body mass index), CRP (c-reactive protein), HOMA-IR (homeostatic model assessment of insulin resistance), ALT (alanine aminotransferase), AST (aspartate aminotransferase), GGT (gamma glutamyl transferase).
